# Modelling the Complex Smoking Exposure History in Assessment of Pan-Cancer Risk

**DOI:** 10.1101/2024.11.07.24316871

**Authors:** Wei Liu, Ya-Ting Chen, Baiwenrui Tao, Ying Lv, Yan-Xi Zhang, Hui-Ying Ren, Yu-Ting Zhang, Yu-Ping Fan, Meng-Han Li, Ya-Xin Shi, Shi-Yuan Wang, Bing-Wei Chen, Frits van Osch, Maurice P. Zeegers, Qi-Rong Qin, Anke Wesselius, Evan Yi-Wen Yu

## Abstract

Modelling complex smoking histories, with more comprehensive and flexible methods, to show what profile of smoking behavior is associated with the risk of different cancers remains poorly understood. This study aims to provide insight into the association between complex smoking exposure history and pan-cancer risk by modelling both smoking intensity and duration in a large-scale prospective cohort. Here, we used data including a total of 0.5 million with cancer incidences of 12 smoking-related cancers. To jointly interpret the effects of intensity and duration of smoking, we modelled excess relative risks (ERRs)/pack-year isolating the intensity effects for fixed total pack-years, thus enabling the smoking risk comparison for total exposure delivered at low intensity (for long duration) and at a high intensity (for short duration). The pattern observed from the ERR model indicated that for a fixed number of pack-years, low intensity/long duration or high intensity/short duration is associated with a different greater increase in cancer risk. Those findings were extended to an increase of time since smoking cessation (TSC) showing a reduction of ERR/pack-year for most cancers. Moreover, individuals with favorable lifestyle behaviors, such as regular physical activity and healthy dietary intakes, were shown to have lower ERRs/pack-year, compared to those with unfavorable lifestyle behaviors. Overall, this study systematically evaluates and demonstrates that for pan-cancer risks, smoking patterns are varied, while reducing exposure history to a single metric such as pack-years was too restrictive. Therefore, cancer screening guidelines should consider detailed smoking patterns, including intensity, duration, and cessation, for more precise prevention strategies.

**Highlights (Key context and significance):**

- Distinct cancer risk patterns emerge based on smoking exposure beyond equal pack-years: smoking duration is a stronger risk factor for some cancers, while smoking intensity dominates for others.
- Time since smoking cessation (TSC) significantly lowers cancer risk: former smokers experience substantial reductions in risk for most cancers within the first 20 years after quitting.
- Favorable lifestyle behaviors mitigate cancer risks: individuals with regular physical activity and healthy diets show lower excess relative risks (ERRs) for most cancers, compared to those with unhealthy habits.
- Tailored cancer screening based on smoking behavior: cancer screening guidelines should consider detailed smoking patterns, including intensity, duration, and cessation, for more precise prevention strategies.

## Introduction

Smoking is an important modifiable risk factor for many cancers but a comprehensive demonstration of its influence by integration of intensity, duration, and cessation has been challenged ^1–3^. While it is helpful to use a single estimate, such as cumulative exposure (e.g., pack-years), to summarize smoking behavior, this approach may not adequately capture the complex relationship between smoking and cancer risk ^4,5^. Although consensus has been reached that modelling pack-years is for the modifying effect of cigarette formulation or ways of targeting screening or other public health interventions, it may fail to reveal subtler phenomena that could shed light on mechanisms ^6^. Several researchers have interpreted pack-years limited in making biologically credible models that provide unbiased information on complex smoking exposure histories ^7,8^ and circumvent multicollinearity issues ^9^. Other time-related modifiers suggest that smoking has multiple effects in the carcinogenic process, thereby, the cooperation of flexible modelling, to capture more complex smoking and enhance the understanding of the molecular basis of smoking-related cancer risk, is needed.

To our knowledge, models that incorporate smoking duration and intensity typically examine these factors in isolation or various combinations, rather than integrating them comprehensively, which can be problematic for exploring complex patterns due to changing total exposures. That is, assessing the smoking risk for either increasing intensity or duration also incorporates the effect of increasing pack-years, leading to biased interpretation and comparison for the independent effect of intensity and duration.

To address all these dimensions together and uncover novel associations, modelling complex smoking exposures has emerged as a powerful tool ^8,10–15^, offering insights that are often missed when examining factors separately, however, evidence specifically linking smoking to pan-cancers remains limited. Although research has long focused on modelling smoking behavior effects, complex smoking exposure—encompassing the simultaneous consideration of smoking duration, intensity, and time since smoking cessation (TSC)—has not been systematically assessed concerning pan-cancers. Additionally, most previous studies have employed a case-control design, which is prone to recall bias and has limited capacity for causal inference. Evidence from large-scale prospective cohort studies remains sparse.

Several studies have suggested that with equal pack-year, individuals who had smoked lower cigarettes/day for a longer duration have different cancer risks compared to those who smoked higher cigarettes/day for a shorter duration ^4,16–19^. These studies have compared excess cancer risks/pack-year across smoking intensity categories, and recent models have incorporated TSC or stratified by age to account for exposure timing ^8,18,20^. Cancer is a complex trait influenced by lifestyle behavior such as smoking, the inconsistency in findings from studies examining the association between cancer risk and complex smoking exposure highlights the need for further research. Therefore, this study aimed to investigate a more nuanced understanding between complex smoking exposure and smoking-related pan-cancer risks, based on our previous study ^19^ that modelled excess cancer risk per pack-year by intensity and TSC continuously and to interpret the various smoking effects, in a large-scale prospective cohort.

## Methods and Materials

### Study Population

Participants for this study were sourced from the UK Biobank, a large-scale prospective study that has been described in detail elsewhere ^21^. In summary, the UK Biobank includes over 500,000 individuals from the United Kingdom. Between 2006 and 2010, invitations were extended to all individuals aged 40 to 69 years living within 25 miles of a study center, from a total of 9.2 million invitations sent. Ultimately, 503,325 participants were recruited. Baseline data collection involved extensive self-reporting through touchscreen tests, questionnaires, and nurse-led interviews, as well as anthropometric assessments and biological sample collection. Health records were supplemented with secondary care data from linked hospital episode statistics (HES).

For our analysis, the UK Biobank database initially included 502,505 participants. We excluded individuals with incomplete or inaccurate smoking status information and data regarding smoking intensity, duration, or pack-years (n=11,431) and those who withdraw with informed consent (n=12), resulting in a final analysis cohort of 491,062 participants (Figure 1). This study utilized the UK Biobank resource under Application #55889.

**Figure 1.**
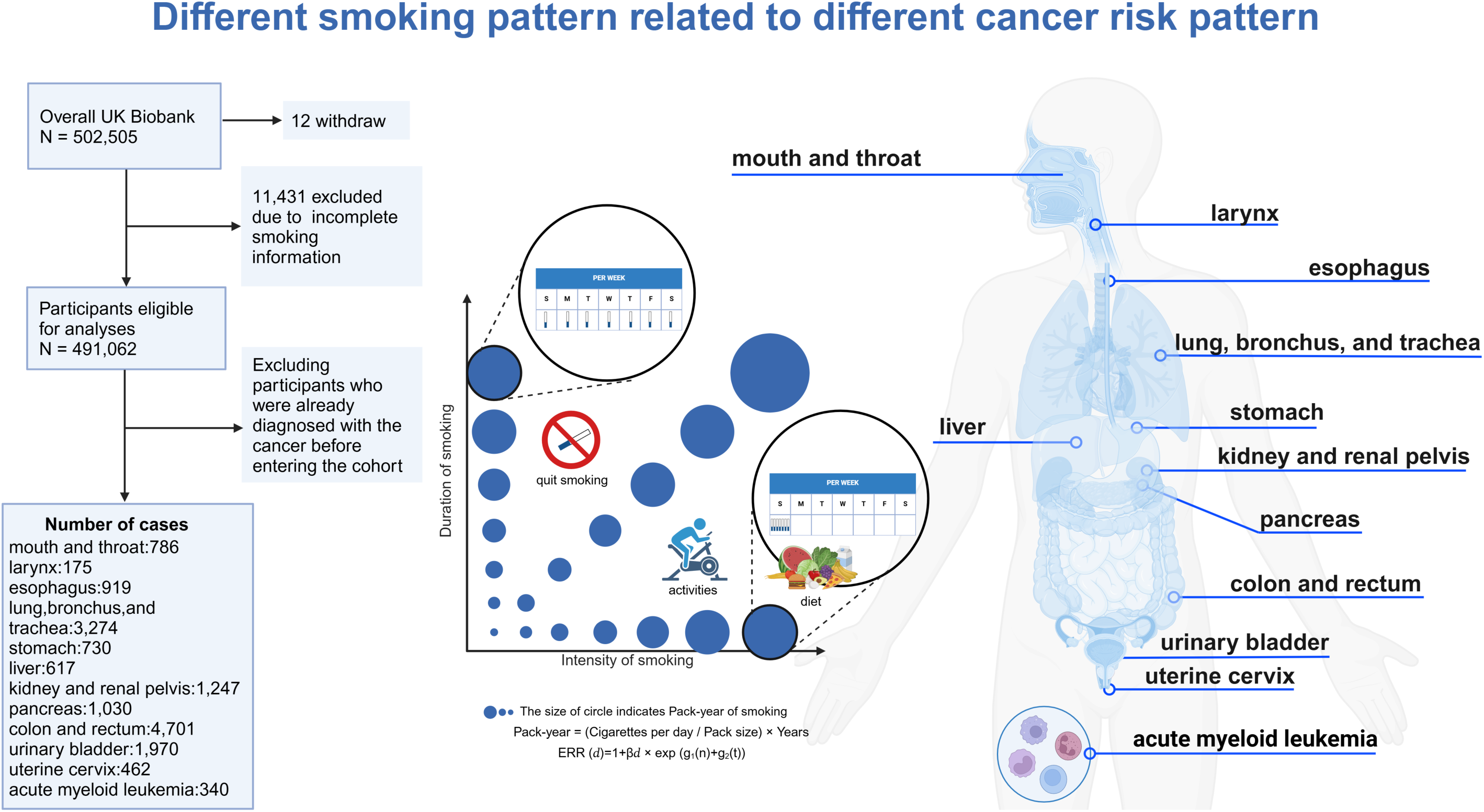
Study overview and workflow This figure illustrates the model formulas and flow chart of participants used in the study, aimed at analyzing the relationship between different smoking patterns and 12 smoking related cancers in the UK Biobank, which includes 491,062 participants into analysis. The diagram in the middle shows two scenarios: high-intensity, low-duration smoking, and low-intensity, high-duration smoking under the same total pack-years. Therefore, we applied a flexible excess relative risk (ERR) model to explore the association of exposure intensity/duration and cancer risk simultaneously. This model isolates the intensity effects while holding total pack-years constant, allowing for the comparison of ERRs for total exposure delivered at lower intensity (for longer duration) and at higher intensity (for shorter duration) across the 12 cancers. Additionally, the model adjusts for the ERR per pack-year after smoking cessation. The figure on the right presents the 12 cancers included in the analysis for which smoking is explicitly identified as a risk factor according to the WHO and US CDC. Abbreviations: ERR, excess relative risk.

### Ascertainment of Pan-Cancers

In this study, “pan-cancers” refer to the 12 cancers associated with smoking as identified by the U.S. CDC ^22^. These include: i) mouth and throat; ii) larynx; iii) esophagus; iv) lung, bronchus, and trachea; v) stomach; vi) liver; vii) kidney and renal pelvis; viii) pancreas; ix) colon and rectum; x) urinary bladder; xi) uterine cervix; and xii) acute myeloid leukemia. Cancer definitions within the UK Biobank are detailed in Table S1 and are based on International Classification of Diseases (ICD) codes (ICD-10 and ICD-9). For each cancer, participants who were diagnosed i) at or before baseline and ii) by self-report were excluded.

### Statistical Analysis and Delivery Rate of Smoking Exposure

We applied a statistical approach based on the models proposed by Vlaanderen et al. ^8^ and Lubin et al. ^18^. Our smoking data included smoking status (never, current, former), pack-years (packs smoked per day × years as a smoker), cigarettes/day, and TSC. Participants were categorized into “never smokers”, “former smokers”, and “current smokers” based on their responses to questions about their smoking history. Those who answered “Yes” to smoking were classified as “current smokers”. Those who answered “No” and indicated a history of smoking were categorized as “former smokers”, while those who neither currently smoked nor had ever smoked were considered “never smokers” ^21^.

We tested for violations of survival time following a piecewise exponential distribution and confirmed that the data adhered to this model ^23^. Consequently, we utilized a Poisson regression model for our analyses. The primary analysis adjusted for age (<45, 45–50, 50–60, 60–65, ≥65 years), sex (men and women), body mass index (BMI, <18.5, 18.5–25, 25–30, ≥30, or unknown, kg/m²), ethnicity (White, Asian or Asian British, Black or Black British, Chinese, Mixed, other ethnic groups, or unknown), alcohol consumption status (never, former, current, or unknown), and socioeconomic status (SES, low, middle, high, or unknown), as main adjustment model. Detailed adjustments are provided in the Supplementary materials. In the UK Biobank, SES was derived from education level, household income, employment status, and the Townsend deprivation index. Education was categorized into various qualifications, while income and employment status were grouped accordingly. The Townsend deprivation index, an area-level SES measure from national census data, was recoded with higher values indicating higher SES ^24^. The overall SES variable was computed by combining these factors and categorized into three groups based on tertiles (low, middle, high, or unknown).

To further examine the impact of lifestyle factors such as physical activity and diet on the relationship between smoking exposure and pan-cancer risk, we adjusted for regular physical activity and healthy dietary scores. Regular physical activity was defined as meeting the 2017 UK Physical Activity Guidelines ^25^. Dietary intake was assessed using the healthy dietary score by Lourida et al., based on the Mediterranean Diet and Heart-Healthy Dietary Priorities ^26,27^. The score included factors such as servings of fruit, vegetables, fish, and meat, and was categorized into healthy (≥4) and unhealthy (<4) diets (Table S2 & S3). We also evaluated potential interactions between TSC and these lifestyle factors ^28,29^ to explore effect modification.

We fitted an exponential model to estimate the excess relative risk (ERRs) per pack-year by smoking intensity to investigate the independent effect of cigarette smoking duration and intensity of cigarette smoking on each cancer risk. In these models, the low intensity with long duration smokers was compared with the high intensity with short duration smokers based on equal pack-years. In addition, TSC was also taken into account, which can expand the scope of the research subjects from current smokers and never smokers to include all former smokers, and incorporating the time after smoking cessation into these models was estimated to better fit the data ^8^.

We used the model as:

RR (d)=1+βd*exp (g_1_(n)+g_2_(t)),

Where β represents the ERR per pack-year at g_1_=1 and g_2_=1, g_1_ and g_2_ are the distribution of lifetime average intensity of cigarette smoking of all smokers and TSC of all former smokers, respectively.

We also selected the optimal transformation of g_1_(n) and g_2_(t) by comparing Akaike information criterion (AIC), with lower AIC indicating a better performance (Table S4). Therefore, we fitted with different g_1_(n) and g_2_(t) transformations per cancer for relatively best performance as: i) G=exp{μ_1_ln(n)+μ_2_ln(t)}; ii) G=exp{μ_1_ln(n)+μ_2_ln(n)^2^+μ_3_ln(t)+μ_4_ ln(t)^2^}; iii) G=exp{μ_1_ln(n)+μ_2_n+μ_3_ln(t)+μ_4_t}; iv) G=exp{μ_1_n+μ_2_n^2^+μ_3_t+μ_4_t^2^} where d represents the pack-years, n represents the cigarettes/day, and t represents the TSC (i.e., years).

The results from such models describe delivery rate patterns of exposure to smoking in relation to pan-cancer risk. The delivery rate is described by estimating how increasing intensity within a fixed number of pack-years influences each cancer risk. For example, an inverse-exposure-rate effect for intensity would mean that the ERR/pack-year (the strength of association) decreases with more cigarettes/day (and thus decreased duration) or the ERR/pack-year increases with lower cigarettes/day (and increased duration). Consequently, for two individuals with equal total pack-years, a greater risk accrues to the individual smoking for lower intensity at a longer duration, or the individual smoking for higher intensity at a shorter duration.

The 95% confidence intervals (CIs) for ERR models were estimated through bootstrapping with 1,000 replications of the original data. To avoid overinterpreting regions with sparse data, we excluded predicted values below the 1st percentile (i.e., <2 cigarettes/day) and above the 99th percentile (i.e., >60 cigarettes/day), applying the same method for TSC predictions.

Sensitivity analyses included: i) different adjustment models; ii) restricting the analysis to participants aged 50–74 years and regrouping former smokers who quit within 5 years as current smokers according to Lubin et al. ^18^; iii) excluding non-White individuals; iv) excluding cancer cases occurring within the first year of follow-up; v) excluding never smokers who reported passive smoking; vi) adjusting for alcohol consumption frequency and volume (available only for current alcohol consumers in UK Biobank).

Statistical analyses were conducted using SAS 9.4 and R software (version 4.0.5). All tests were two-sided, with *p* values <0.05 considered statistically significant.

## Results

### Population Characteristics

Table 1 summarizes smoking data for 12 types of cancer in the UK Biobank, categorized by sex. Compared to non-cancer controls, individuals with cancer exhibited worse smoking behaviors across several metrics, including the proportion of ever smokers and current smokers, duration, intensity, and pack-years. This suggests that smoking is a significant risk factor for cancer development. This study includes 491,062 study participants with a median follow-up of 4.93 years for cancer cases and 11.63 years for non-cancer controls. Among cancer cases, 42.36% were women, while 55.01% participants in the overall cohort were women. The mean (SD) age at recruitment was 61.34 (6.64) years for cancer cases and 56.85 (8.10) for non-cancer controls (Table S5).

**Table 1.**
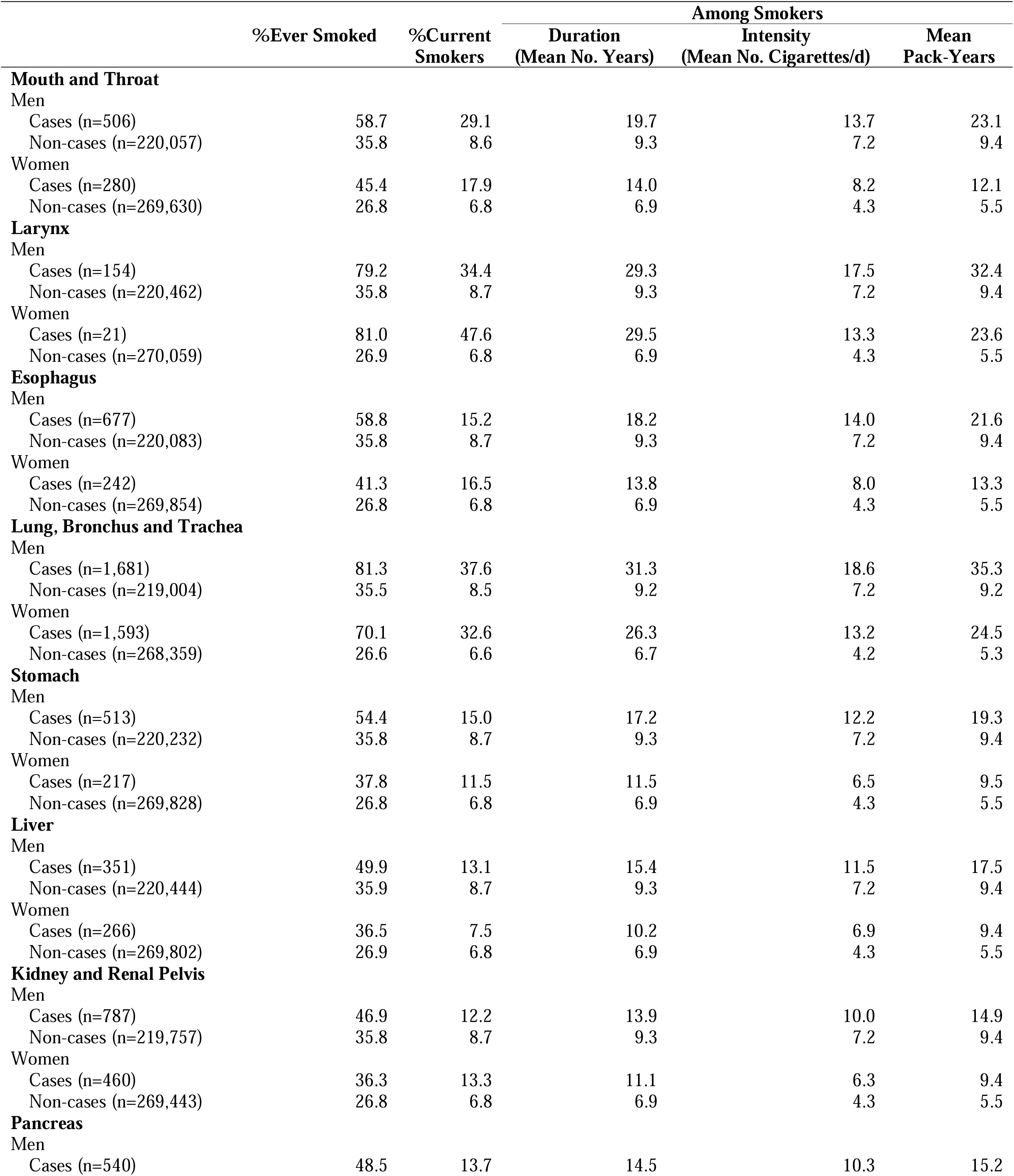
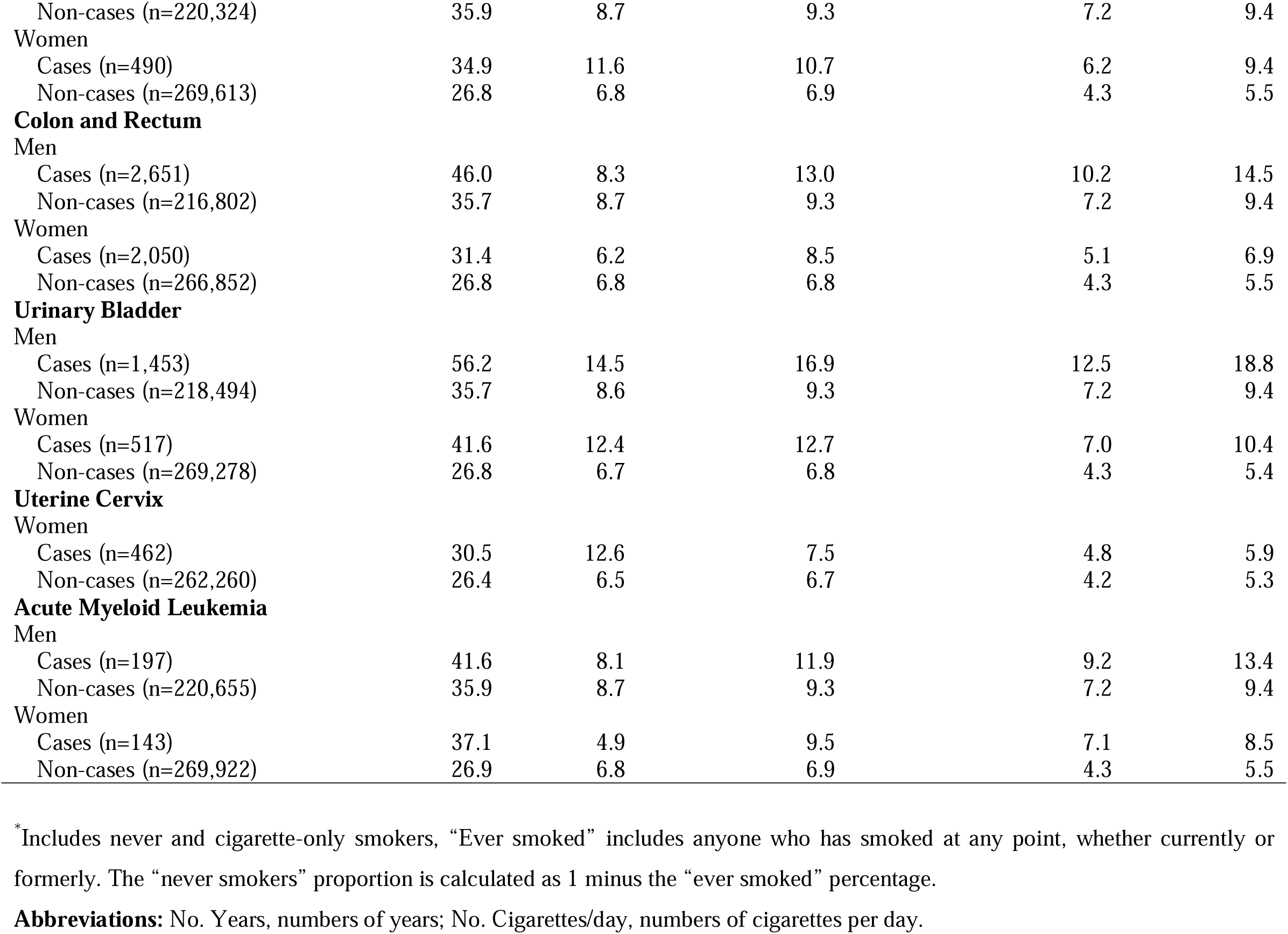
Summary of smoking data* for 12 cancers in the UK Biobank by sex.

|  | %Ever Smoked | %Current Smokers | Among Smokers |  |  |
| --- | --- | --- | --- | --- | --- |
|  |  |  | Duration<br>(Mean No. Years) | Intensity<br>(Mean No. Cigarettes/d) | Mean<br>Pack-Years |
| **Mouth and Throat** |  |  |  |  |  |
| Men |  |  |  |  |  |
| Cases (n=506) | 58.7 | 29.1 | 19.7 | 13.7 | 23.1 |
| Non-cases (n=220,057) | 35.8 | 8.6 | 9.3 | 7.2 | 9.4 |
| Women |  |  |  |  |  |
| Cases (n=280) | 45.4 | 17.9 | 14.0 | 8.2 | 12.1 |
| Non-cases (n=269,630) | 26.8 | 6.8 | 6.9 | 4.3 | 5.5 |
| **Larynx** |  |  |  |  |  |
| Men |  |  |  |  |  |
| Cases (n=154) | 79.2 | 34.4 | 29.3 | 17.5 | 32.4 |
| Non-cases (n=220,462) | 35.8 | 8.7 | 9.3 | 7.2 | 9.4 |
| Women |  |  |  |  |  |
| Cases (n=21) | 81.0 | 47.6 | 29.5 | 13.3 | 23.6 |
| Non-cases (n=270,059) | 26.9 | 6.8 | 6.9 | 4.3 | 5.5 |
| **Esophagus** |  |  |  |  |  |
| Men |  |  |  |  |  |
| Cases (n=677) | 58.8 | 15.2 | 18.2 | 14.0 | 21.6 |
| Non-cases (n=220,083) | 35.8 | 8.7 | 9.3 | 7.2 | 9.4 |
| Women |  |  |  |  |  |
| Cases (n=242) | 41.3 | 16.5 | 13.8 | 8.0 | 13.3 |
| Non-cases (n=269,854) | 26.8 | 6.8 | 6.9 | 4.3 | 5.5 |
| **Lung, Bronchus and Trachea** |  |  |  |  |  |
| Men |  |  |  |  |  |
| Cases (n=1,681) | 81.3 | 37.6 | 31.3 | 18.6 | 35.3 |
| Non-cases (n=219,004) | 35.5 | 8.5 | 9.2 | 7.2 | 9.2 |
| Women |  |  |  |  |  |
| Cases (n=1,593) | 70.1 | 32.6 | 26.3 | 13.2 | 24.5 |
| Non-cases (n=268,359) | 26.6 | 6.6 | 6.7 | 4.2 | 5.3 |
| **Stomach** |  |  |  |  |  |
| Men |  |  |  |  |  |
| Cases (n=513) | 54.4 | 15.0 | 17.2 | 12.2 | 19.3 |
| Non-cases (n=220,232) | 35.8 | 8.7 | 9.3 | 7.2 | 9.4 |
| Women |  |  |  |  |  |
| Cases (n=217) | 37.8 | 11.5 | 11.5 | 6.5 | 9.5 |
| Non-cases (n=269,828) | 26.8 | 6.8 | 6.9 | 4.3 | 5.5 |
| **Liver** |  |  |  |  |  |
| Men |  |  |  |  |  |
| Cases (n=351) | 49.9 | 13.1 | 15.4 | 11.5 | 17.5 |
| Non-cases (n=220,444) | 35.9 | 8.7 | 9.3 | 7.2 | 9.4 |
| Women |  |  |  |  |  |
| Cases (n=266) | 36.5 | 7.5 | 10.2 | 6.9 | 9.4 |
| Non-cases (n=269,802) | 26.9 | 6.8 | 6.9 | 4.3 | 5.5 |
| **Kidney and Renal Pelvis** |  |  |  |  |  |
| Men |  |  |  |  |  |
| Cases (n=787) | 46.9 | 12.2 | 13.9 | 10.0 | 14.9 |
| Non-cases (n=219,757) | 35.8 | 8.7 | 9.3 | 7.2 | 9.4 |
| Women |  |  |  |  |  |
| Cases (n=460) | 36.3 | 13.3 | 11.1 | 6.3 | 9.4 |
| Non-cases (n=269,443) | 26.8 | 6.8 | 6.9 | 4.3 | 5.5 |
| **Pancreas** |  |  |  |  |  |
| Men |  |  |  |  |  |
| Cases (n=540) | 48.5 | 13.7 | 14.5 | 10.3 | 15.2 |
| Non-cases (n=220,324) | 35.9 | 8.7 | 9.3 | 7.2 | 9.4 |
| **Women** |  |  |  |  |  |
| Cases (n=490) | 34.9 | 11.6 | 10.7 | 6.2 | 9.4 |
| Non-cases (n=269,613) | 26.8 | 6.8 | 6.9 | 4.3 | 5.5 |
| **Colon and Rectum** |  |  |  |  |  |
| **Men** |  |  |  |  |  |
| Cases (n=2,651) | 46.0 | 8.3 | 13.0 | 10.2 | 14.5 |
| Non-cases (n=216,802) | 35.7 | 8.7 | 9.3 | 7.2 | 9.4 |
| **Women** |  |  |  |  |  |
| Cases (n=2,050) | 31.4 | 6.2 | 8.5 | 5.1 | 6.9 |
| Non-cases (n=266,852) | 26.8 | 6.8 | 6.8 | 4.3 | 5.5 |
| **Urinary Bladder** |  |  |  |  |  |
| **Men** |  |  |  |  |  |
| Cases (n=1,453) | 56.2 | 14.5 | 16.9 | 12.5 | 18.8 |
| Non-cases (n=218,494) | 35.7 | 8.6 | 9.3 | 7.2 | 9.4 |
| **Women** |  |  |  |  |  |
| Cases (n=517) | 41.6 | 12.4 | 12.7 | 7.0 | 10.4 |
| Non-cases (n=269,278) | 26.8 | 6.7 | 6.8 | 4.3 | 5.4 |
| **Uterine Cervix** |  |  |  |  |  |
| **Women** |  |  |  |  |  |
| Cases (n=462) | 30.5 | 12.6 | 7.5 | 4.8 | 5.9 |
| Non-cases (n=262,260) | 26.4 | 6.5 | 6.7 | 4.2 | 5.3 |
| **Acute Myeloid Leukemia** |  |  |  |  |  |
| **Men** |  |  |  |  |  |
| Cases (n=197) | 41.6 | 8.1 | 11.9 | 9.2 | 13.4 |
| Non-cases (n=220,655) | 35.9 | 8.7 | 9.3 | 7.2 | 9.4 |
| **Women** |  |  |  |  |  |
| Cases (n=143) | 37.1 | 4.9 | 9.5 | 7.1 | 8.5 |
| Non-cases (n=269,922) | 26.9 | 6.8 | 6.9 | 4.3 | 5.5 |

\*Includes never and cigarette-only smokers, “Ever smoked” includes anyone who has smoked at any point, whether currently or formerly. The “never smokers” proportion is calculated as 1 minus the “ever smoked” percentage.
**Abbreviations:** No. Years, numbers of years; No. Cigarettes/day, numbers of cigarettes per day.

Among current smokers with cancer, 1,878 (75.00%) smoked more than 10 cigarettes/day and 2,410 (96.25%) had a smoking duration longer than 20 years. This contrasts with non-cancer controls, who 22,363 (64.01%) smoked more than 10 cigarettes/day and 32,780 (93.83%) had a smoking duration longer than 20 years. For former smokers with cancer, 4,304 (82.31%) smoked more than 10 cigarettes/day, and 3,499 (66.92%) had a smoking duration longer than 20 years. Additionally, 4,283 participants (81.91%) had a TSC greater than 5 years, compared to 92,874 participants (85.05%) in the non-cancer control group, respectively (Table S6).

### Complex Smoking Exposure in Relation to Different Patterns of Pan-cancer Risk

Figure 2 illustrates the ERR/pack-year and 95% CI for various smoking intensities, considering smoking cessation (based on the main adjustment model). The curve shows different smoking delivery rate patterns with different cancer risks; as daily cigarette consumption increases (with reduced smoking duration), the ERRs/pack-year exhibit varying changes with either an increased pattern or a decreased pattern. This indicates that for equal pack-years, smoking for a longer duration (at lower cigarettes/day) is differentially associated with different cancer risks than smoking higher cigarettes/day (at a shorter duration). Specifically, we observed the lower cigarettes/day over a longer duration showing a greater ERR/pack-year of cancers than smoking more cigarettes/day over a shorter duration (e.g., cancer sites of the stomach, urinary bladder, pancreas, and kidney), while the higher cigarettes/day over a shorter duration showing a greater ERR/pack-year of cancers than smoking lower cigarettes/day over a longer duration (e.g., cancer sites of mouth and throat, esophagus, liver, uterine cervix, and acute myeloid leukemia). In cancer sites of the lung, bronchus and trachea, larynx, and colon and rectum, we observed an overall decreased ERRs/pack-year of smoking pattern with cigarettes/day increment, while an altered point at 40 cigarettes/day with slightly increased ERRs/pack-year of smoking pattern after. The risk estimate remained essentially unchanged regardless of which other covariates were included in the models (Figure S1–S5).

**Figure 2.**
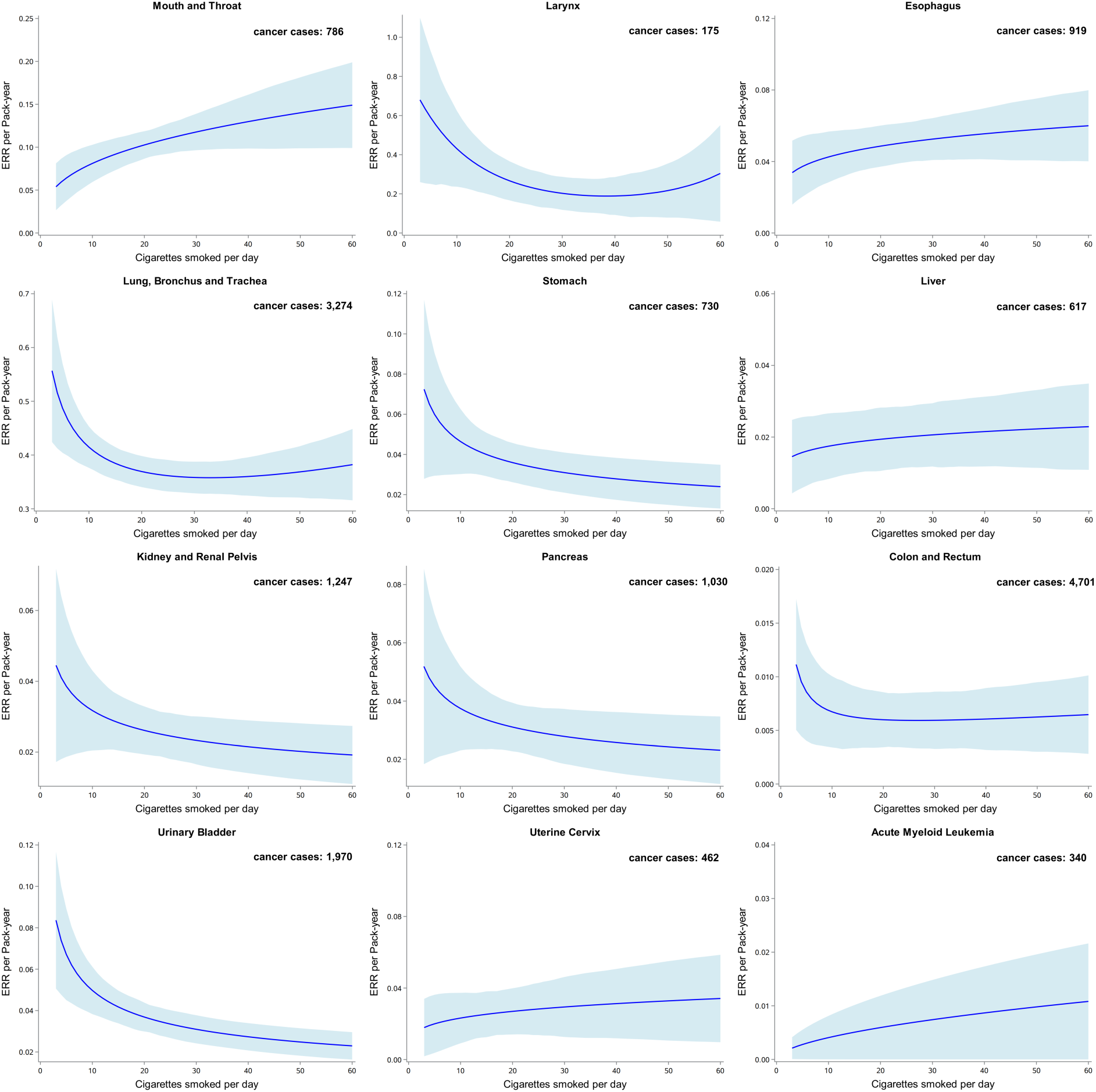
The excess relative risk (ERR) for pan-cancers per pack-year of smoking by smoking intensity. Bootstrapped 95% confidence intervals are based on 1,000 replications. The blue solid line represents the prediction from the function g1(n) in the model, with the area showing 95% confidence interval obtained from bootstrap analysis. Model adjustments: age (<45, 45–50, 50–60, 60–65, ≥65, years), sex (men or women), ethnicity (White, Asian or Asian British, Black or Black British, Chinese, Mixed, other ethnic group, or unknown), BMI (<18.5, 18.5–25, 25–30, ≥30, or unknown, kg/m2), socioeconomic status (low, medium, high, or unknown), and alcohol consumption status (never, previous, current, or unknown). Abbreviations: ERR, excess relative risk; BMI, body mass index.

By comparing the mean ERRs/pack-year, we found the association strength was varied across cancers, where some of them yielded an ERR/pack-year over 10% (e.g., cancer sites of lung, bronchus and trachea, larynx, and mouth) but others with an ERR/pack-year under 10% for lifetime smoking behavior when accounting for duration and intensity together (Figure S6).

### Excess Risk of Pan-Cancer in Relation to Time Since Smoking Cessation (TSC)

We examined the association of smoking cessation with the ERR/pack-year for pan-cancers considering cumulative pack-years smoked, in an attempt to control for confounding by pack-years. After partitioning of the qualitative effect of former smoking, TSC was associated with significant risk reductions for 10 cancers, mostly reaching a relatively small ERR/pack-year in the first 20 years, in comparison with current and never smokers (Figure 3). Again, the risk estimate remained essentially unchanged regardless of which other dimensions of smoking or covariates were included in the models (Figure S7–S11). By comparing the Min-Max ranges of the ERR/pack-year estimates, we found for the cancers showing decreased ERR/pack-year with TSC, the majority of them yielded a ratio of range of excess risk/maximum ERR over 50%, indicating quitting smoking could significantly reduce the ERRs when considering a more complex smoking exposure (Table S8).

**Figure 3.**
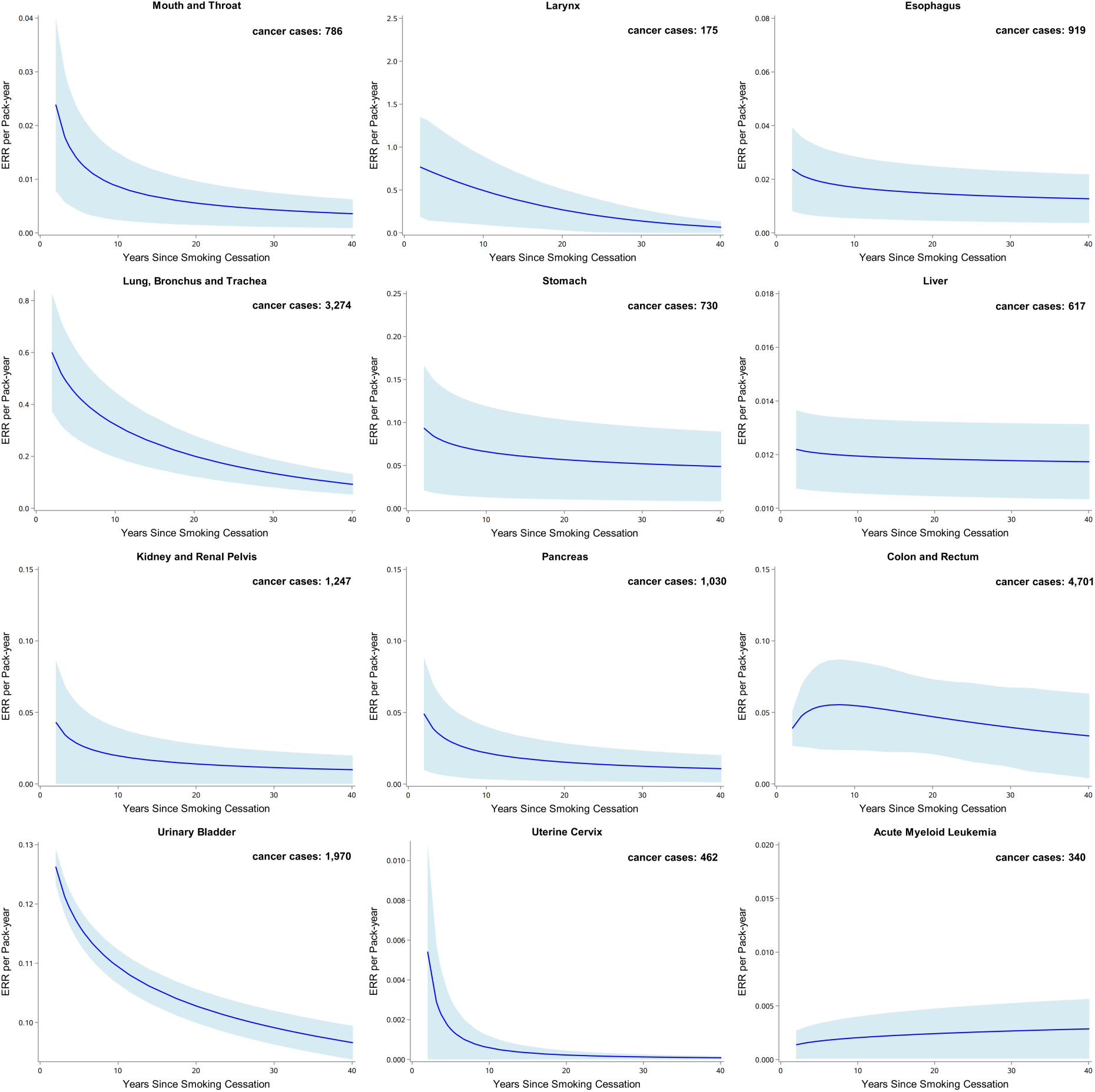
The excess relative risk (ERR) for pan-cancers per pack-year of smoking by time since smoking cessation. Bootstrapped 95% confidence intervals are based on 1,000 replications. The blue solid line represents the prediction from the function g2(t) in the model, with the area showing 95% confidence interval obtained from bootstrap analysis. Model adjustments: age (<45, 45–50, 50–60, 60–65, ≥65, years), sex (men or women), ethnicity (White, Asian or Asian British, Black or Black British, Chinese, Mixed, other ethnic group, or unknown), BMI (<18.5, 18.5–25, 25–30, ≥30, or unknown, kg/m2), socioeconomic status (low, medium, high, or unknown), and alcohol consumption status (never, previous, current, or unknown). Abbreviations: ERR, excess relative risk; BMI, body mass index.

However, we observed an inconsistent change of TSC pattern with ERRs/pack-year for colon and rectum cancer, the ERR/pack-year was increased within 7 years but decreased after; and for acute myeloid leukemia, the ERRs/pack-year still increased. Those findings suggest for some cancers, the reduction of cancer risk at the beginning of smoking cessation was temporal with a possible forward increased risk, which could be biased by other residual factors or due to statistical power. Particularly for colon and rectum cancer in line with a previous study that observed the cancer risk seems to increase for approximately 7 years after quitting smoking, before gradually decreasing ^30^. This may reflect that, in the early stages of smoking cessation, the health effects caused by prior smoking persist, which is especially true for individuals who have already sustained severe damage from smoking, referred to as “sick quitters”. Even after quitting, the damage may lead to a higher short-term cancer risk. As for acute myeloid leukemia, we hypothesized the harmful effect of smoking on the increase of cancer risk might not alleviate or diminish once quitting smoking, which is partially supported by a review showing quitting smoking did not stop the increased risks of acute myeloid leukemia at once ^31^.

### Effect Modification by Major Lifestyle Factors

In recent years, smoking, physical activity, and diet have been recognized as key lifestyle factors influencing human health, often contributing to the formulation of a healthy lifestyle score. In this study, we examined the impact of physical activity and diet—both separately and in combination—on the relationship between complex smoking exposure and pan-cancer risk. Our analysis revealed that neither physical activity nor diet significantly altered the associations between smoking intensity-duration patterns, TSC, and ERR/pack-year for various cancers (see Figure S12–S16 for smoking intensity and Figure S17–S21 for TSC).

However, given the modifiable impact of physical activity on smoking cessation, we further compared smoking patterns across stratified analyses of regular versus irregular physical activity. The patterns remained largely consistent with those observed in the main adjustment model, but we noted that for most cancers, ERR/pack-year with TSC was lower among individuals engaging in regular physical activity (Figure 4). Similarly, when stratifying by the healthy dietary score—healthy versus unhealthy—we found that smoking patterns were comparable to those in the main adjustment model. Notably, a healthy dietary score was associated with lower ERRs with increasing TSC compared to an unhealthy dietary score for most cancers (Figure 5). The differences between favorable and unfavorable lifestyle behaviors were statistically significant (*p* < 0.05) as shown in Table S7. These findings suggest that maintaining a healthy lifestyle significantly modifies the relationship between smoking and cancer risk. While most cancers showed reduced ERRs/pack-year with favorable lifestyle behaviors, a few cancers exhibited either similar or partially reduced ERRs/pack-year among those with less favorable behaviors.

**Figure 4.**
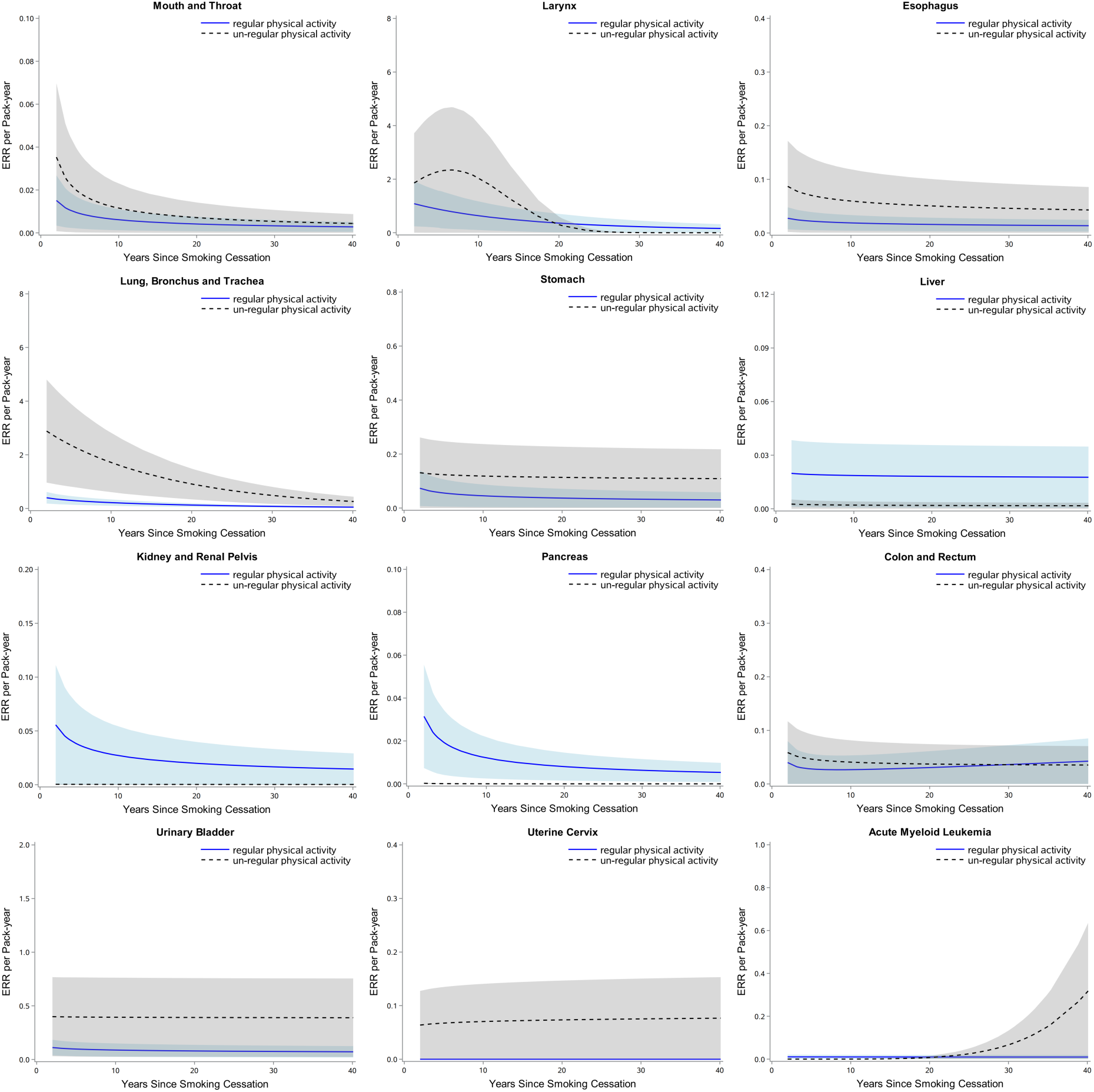
The excess relative risk (ERR) for pan-cancers per pack-year of smoking by time since smoking cessation within categories of physical activity. Bootstrapped 95% confidence intervals are based on 1,000 replications. The blue solid line represents the prediction from the function g2(t) in the model for individuals who engage in regular physical activity, with the area showing 95% confidence interval obtained from bootstrap analysis. Model adjustments: age (<45, 45–50, 50–60, 60–65, ≥65, years), sex (men or women), ethnicity (White, Asian or Asian British, Black or Black British, Chinese, Mixed, other ethnic group, or unknown), BMI (<18.5, 18.5–25, 25–30, ≥30, or unknown, kg/m2), socioeconomic status (low, medium, high, or unknown), alcohol consumption status (never, previous, current, or unknown). Abbreviations: ERR, excess relative risk; BMI, body mass index.

**Figure 5.**
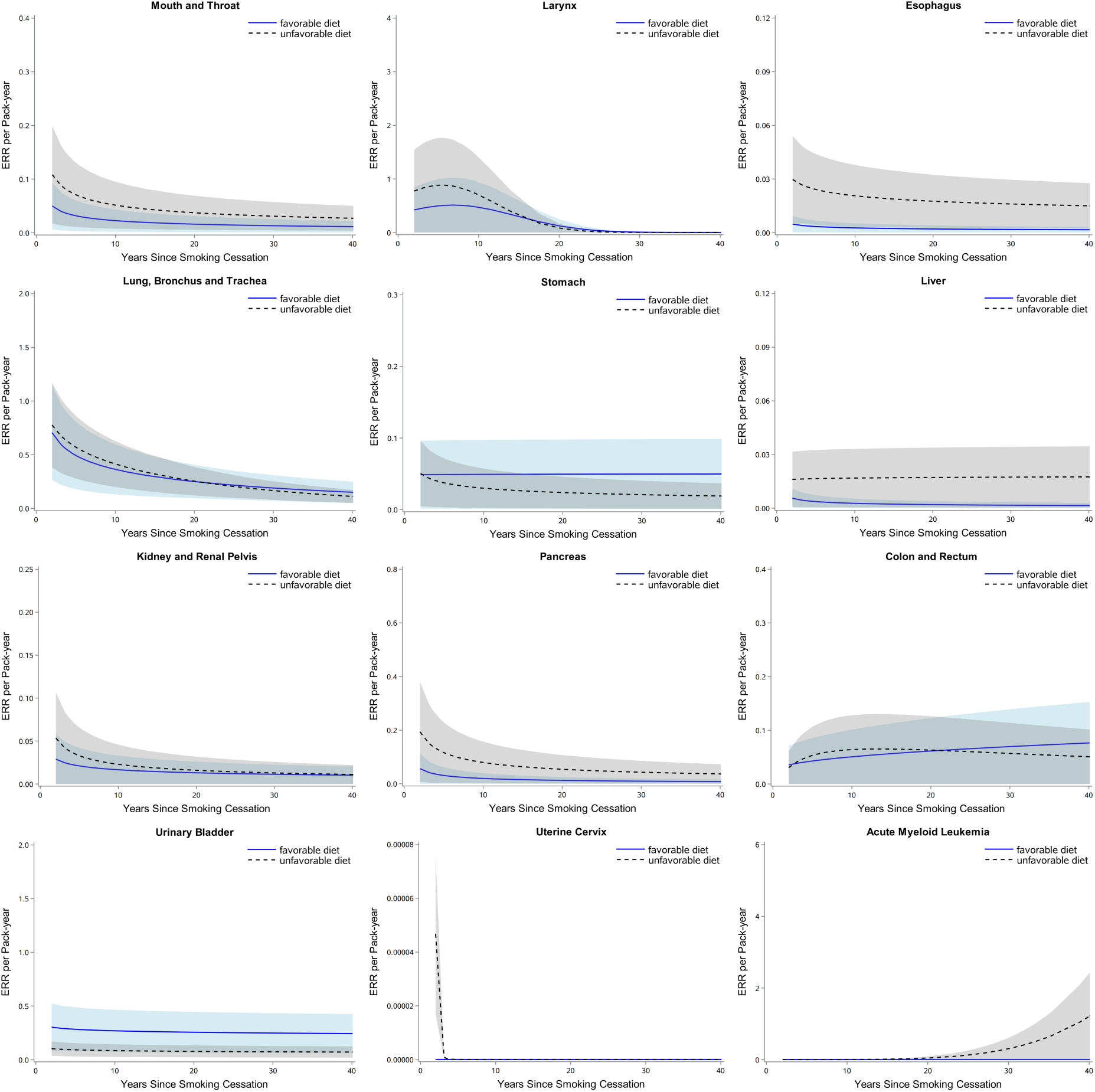
The excess relative risk (ERR) for pan-cancers per pack-year of smoking by time since smoking cessation within categories of a healthy diet. Bootstrapped 95% confidence intervals are based on 1,000 replications. The blue solid line represents the prediction from the function g2(t) in the model for individuals who engage in a healthy diet, while the dashed black line corresponds to those with an unhealthy diet. Model adjustments: age (<45, 45–50, 50–60, 60–65, ≥ 65, years), sex (men or women), ethnicity (White, Asian or Asian British, Black or Black British, Chinese, Mixed, other ethnic group, or unknown), BMI (<18.5, 18.5–25, 25–30, ≥30, or unknown, kg/m2), socioeconomic status (low, medium, high, or unknown), alcohol consumption status (never, previous, current, or unknown). Abbreviations: ERR, excess relative risk; BMI, body mass index.

### Sensitivity Analysis

Wide confidence intervals observed in some patterns indicate that certain cancers or cigarette consumption levels had limited statistical power, which may account for the unexpected and inconsistent trends in ERRs/pack-year for some cancers. These confidence intervals should be interpreted with caution. For many cancers where a pattern could be discerned, there was considerable uncertainty in effect modification by smoking intensity per pack-year, particularly at very low or very high extremes, despite excluding <1% and >99% of extreme intensity values. Additionally, the cumulative effect of heavy smoking, such as exceeding 40 cigarettes/day, should be considered, as there might be a potential plateau effect in smoking risk^32^. The bootstrapped 95% CIs show that the most reliable data is for individuals smoking between 10 and 40 cigarettes/day, as this range included the majority of smokers in the UK Biobank, making the shape of the curve most dependable within this interval.

Sensitivity analyses, incorporating physical activity and diet into the fully adjusted model, yielded consistent results regarding the association between complex smoking exposure and pan-cancer risks for both intensity-duration combinations per fixed pack-year and TSC (see Figure S12–S16 for smoking intensity and Figure S17–S21 for TSC). Comparisons across different adjusted models showed that the range of ERR estimates and patterns was consistent for most cancer sites, except for uterine cervix cancer and liver. This consistency reinforces the validity of our flexible ERR model ^32^.

## Discussion

### Complexity of Smoking Exposure and Cancer Risk

Our study explored the intricate relationships between smoking exposure—encompassing duration, intensity, and time since cessation (TSC)—and cancer risk across various cancer types. We observed that different smoking patterns are associated with distinct cancer risks. Specifically, smoking lower cigarettes/day over a prolonged period was linked to a higher ERR/pack-year for cancers such as the stomach, urinary bladder, pancreas, and kidney. Conversely, higher daily smoking intensity over a shorter duration was more strongly associated with cancers like mouth and throat, esophagus, liver, uterine cervix, and acute myeloid leukemia. Furthermore, the impact of TSC on cancer risk varied: generally, longer cessation periods were associated with reduced risk, though some exceptions were noted. These findings underscore the complex interplay between smoking exposure patterns, cessation, and cancer risk.

### Challenges in Measuring Smoking Exposure and the Role of Duration

Accurately measuring smoking intensity presents significant challenges due to fluctuations in the number of cigarettes smoked/day, variability in cigarette formulations, and differences in smoking behavior. Factors such as the depth of inhalation also influence exposure assessments, leading to potential misclassification biases and constrained reporting ranges, which can result in underestimation or overestimation of risk.

These challenges are further complicated by the need to understand how smoking intensity, beyond merely counting cigarettes, affects carcinogenic processes through various molecular and biological pathways ^33^. Increasing smoking intensity may activate or alter specific molecular mechanisms that influence cancer initiation and progression. Despite substantial research into smoking-induced carcinogenesis, the precise mechanisms linking complex exposure patterns—such as duration, intensity, and TSC—remain inadequately explored.

Current models that incorporate smoking duration, intensity, and TSC, while accounting for pack-years, offer a more comprehensive understanding of smoking’s multifaceted nature.

Our findings highlight the critical role of smoking duration in cancer risk. Extended smoking periods are associated with a greater accumulation of genetic and epigenetic changes ^34,35^, particularly for cancers of the urinary bladder, kidney, pancreas, and stomach. While personal monitoring and biochemical assays provide valuable insights into smoking burden ^33^, they face limitations related to feasibility and cost, especially for cancers with long latency periods. Reviews indicate that smoking-related cancer risks involve complex mechanisms, including nicotine stimulation, increased inflammation, and tobacco smoke particulates ^36^, emphasizing the need for comprehensive exposure models to fully elucidate the impact of smoking duration, intensity, and overall exposure.

### The Role of Smoking Duration and Age in Cancer Risk

Our study, consistent with other research (Table S9), confirms that smoking duration is a key determinant of cancer risk, particularly due to its nonlinear effects on long-term exposure to tobacco carcinogens. Longer smoking durations are associated with increased accumulation of genetic and epigenetic changes, which heighten cancer risk ^34,35^. The role of age in carcinogenesis, particularly age at smoking onset, complicates the analysis of smoking duration’s effects. Smoking duration and age of onset are strongly correlated (Spearman’s correlation test *p* <0.001), and adjusting for age at enrolment and age of smoking onset did not significantly alter the relationship between smoking duration and cancer risk (data not shown). Moreover, the model incorporating smoking duration and age onset of smoking might cause colinear and overfitting, therefore, the close correlation of age at initiation, attained age, and the duration of smoking effectively prevents us from studying the effects of age at initiation in this study ^9,20^. This suggests that while age at initiation and smoking duration are interrelated, their independent effects are challenging to disentangle.

Our findings reinforce the importance of smoking duration as a critical factor in cancer risk and support public health initiatives aimed at delaying smoking initiation among adolescents. Measures such as excise taxes and counter-advertising are crucial, though their effectiveness is limited by cancer’s long latency period and the challenges in measuring exposure time ^20^. Therefore, more effective smoking cessation programs are needed to encourage early quitting.

The inclusion of TSC in our study further emphasizes that quitting smoking reduces cancer risk, highlighting that it is never too late to quit. TSC is highly sensitive to smoking duration ^37^, underscoring the need to consider both duration and intensity in cancer risk assessments. While pack-years—combining cigarettes/day and smoking duration—provide a measure of total exposure, the relative contributions of smoking intensity versus duration to cancer development are not fully understood. Our study suggests that smoking duration is more reliably recalled and measured than daily cigarette intake, which is subject to fluctuation and less accurately quantified.

### Smoking Intensity, Time Since Cessation (TSC), and Cancer Risk

Our study underscores the significant impact of smoking intensity on cancer risk, particularly within the range of 10–40 cigarettes/day. This relationship is largely consistent across various cancer sites, reflecting how smoking intensity influences site-specific disease risk. Tobacco metabolites, including nicotine and cotinine, may contribute to local immunosuppressive effects and DNA damage, potentially explaining these patterns ^8,38^. Increased ERRs/pack-year with higher smoking intensity could result from mechanisms such as metabolic saturation or enhanced DNA repair capacity ^8,38^.

However, the relationship between smoking intensity and cancer risk is complex. High-intensity smoking (e.g., 40 or more cigarettes/day) is associated with higher risks for cancers such as those of the larynx and lung, but this relationship may diminish at very high intensities due to misclassification or variations in inhalation patterns. Our study found that after excluding extreme values, intensity effects remained consistent but showed an increase above 40 cigarettes/day. Results for very low and high intensities should be interpreted with caution, necessitating further research.

TSC also plays a critical role in modifying cancer risk. Longer cessation periods generally reduce cancer risk, although exceptions exist. For example, the ERRs/pack-year of colon and rectal cancer increases within the first 7 years of quitting but may decrease afterward, while ERRs/pack-year of acute myeloid leukemia initially rises with longer cessation. This suggests that TSC interacts with other factors like age and genetic predisposition, affecting the overall cancer risk profile.

Previous research indicates that TSC is a major driver of cancer mortality, with varying sensitivity to smoking duration across different cancers ^8,39^. For instance, lung cancer risk is more strongly associated with smoking duration than with average daily cigarette consumption. Our study highlights that TSC, in conjunction with smoking duration, significantly influences cancer risk profiles and has implications for tobacco cessation strategies. The pack-years measure, which combines cigarettes/day and smoking duration, may not fully capture the differential impacts of these factors on cancer risk.

Our analysis suggests that smoking intensity impacts cancer risk through biological processes such as reduced DNA repair capacity at lower intensities or increased detoxification enzyme induction at higher intensities ^4^. However, patterns of intensity effects may also reflect nicotine satiation, where carcinogenic yield per cigarette decreases as smokers increase their daily intake to maintain addiction-sufficient nicotine levels. This makes the number of cigarettes/day an overestimate of internal exposure rates.

The use of pack-years has been criticized for assuming equal importance of smoking intensity and duration in determining cancer risk. Our study demonstrates that smoking duration or intensity alone may be more strongly associated with cancer risk than the other. Relying solely on pack-years for screening may exclude high-risk individuals who smoke less intensely. Thus, a paradigm shift is needed in assessing and reporting smoking history, incorporating both smoking duration and intensity into more complex exposure models for cancer susceptibility screening.

Additionally, our study explored interactions between smoking and other factors such as physical activity and diet. These interactions significantly modify the association between smoking and cancer risk, emphasizing the need for integrated models that account for these variables.

### Strengths and Limitations of the Excess Relative Risk (ERR) Model

We applied a model similar to the ERR model described by Vlaanderen et al. ^8^, building on earlier approaches such as the Lubin et al. ^18^ model. Our model effectively addresses differences in risk between low intensity/long duration and high intensity/short duration smokers with equal pack-years.

Major strengths of our study include a large-sample-size and well-characterized cohort of current and former smokers with diverse ethnicity and sex, and valid information on smoking and potential confounding factors. The prospective design minimized recall bias and allowed us to examine multiple smoking exposures within the same cohort. Our consistent findings across different cancer types, with multiple adjustments and sensitivity analyses, strengthen our inferences and reduce the likelihood of chance or differential reporting.

However, our study also has limitations. While the ERR model provides detailed insights, some factors remain unaccounted for. For example, more vigorous inhalation patterns, often associated with lighter smokers, could confound risk estimates between heavy and light smokers. Discrepancies between self-reported and objective information were more likely among long-term heavy smokers, potentially leading to inaccurate reporting of intensity and duration. Variations in smoking intensity over time and by age of exposure, along with reliance on self-reported averages, may introduce errors in exposure measurements. To mitigate information bias and residual confounding, we excluded extreme values of cigarette intensity and duration from our analysis. Due to some missing data and the criteria for the definition of favorable/unfavorable on physical activity and dietary intakes, the stratified findings and interpretations upon the impact of lifestyle behaviors on excess risks when quitting smoking should be with caution. Even though competing risks could influence results since smokers were more likely to be selectively removed from follow-up due to other diseases, a previous study ^14^ has demonstrated when assessing complex smoking history, competing risks had no appreciable effect on the results which suggested that any potential bias from competing risk considerations was minimal. Finally, the observational nature of our study limits our ability to uncover molecular or biological pathways linking smoking exposure to cancer development, though it highlights important associations that warrant further research.

## Conclusions

In summary, our study reveals qualitative and quantitative differences in the association between dimensions of smoking exposure and cancer risk. Smoking duration was a strong determinant of cancers such as the urinary bladder, kidney, pancreas, and stomach, while intensity was a major determinant for cancers including the esophagus, liver, and acute myeloid leukemia, with significant nonlinear dose effects observed. TSC emerged as an independent and dominant predictor for most cancers, emphasizing the substantial benefits of quitting smoking in reducing cancer risk, regardless of the duration or intensity of smoking.

## Ethics Approval

The studies involving human participants were reviewed and approved by NHS National Research Ethics Service North West (11/NW/0382). The patients/participants provided their written informed consent to participate in this study.

## Author Contributions

Conceptualization, E.Y.W.Y. and A.W.; Data curation, W.L., Y.C. and B.T.; Formal analysis, W.L. and Y.C.; Funding acquisition, E.Y.W.Y. and Q.Q.; Investigation, W.L. and Y.C.; Methodology, W.L., Y.C. and E.Y.W.Y.; Project administration, E.Y.W.Y. and A.W.; Resources, E.Y.W.Y. and A.W.; Supervision, E.Y.W.Y. and A.W.; Writing-original draft, W.L., A.W. and E.Y.W.Y.; and all other authors Y.L., Y.X.Z., H.Y.R., Y.T.Z., Y.P.F., M.H.L., Y.X.S., S.Y.W., B.W.C., F.O., MP.Z. Q.Q. reviewed, edited the writing, and approved the final manuscript.

## Data Availability

All data produced are available online at UK Biobank.

## Acknowledgments

This study was supported by the: National Natural Science Foundation of China (NSFC, 82204033); the Natural Science Foundation of Jiangsu grant (BK2022020826); Fundamental Research Funds for the Central Universities of China (2242022R10062/3225002202A1), Medical Foundation of Southeast University (4060692202/021), Zhishan Young Scholar Award at the Southeast University (2242023R40031); The Scientific Research Project for Health Commission of Anhui Province (AHWJ2023A20172; AHWJ2023BAa20055). The funders had no role in the study design, data collection, decision to publish, or preparation of the manuscript. The authors acknowledge certain figures (Figure 1) were created, adapted, and exported from BioRender.com (2024). Retrieved from https://app.biorender.com/biorender-templates.

## Data and Code Availability Statement

This work has been conducted using the UK Biobank Resource under Application Number 55889. The UK Biobank is an open-access resource and bona fide researchers can apply to use the UK Biobank dataset by registering and applying at http://ukbiobank.ac.uk/register-apply/. Further information and the key codes for analysis in this study are available from the corresponding author upon request.

## Abbreviations

ERR, excess relative risk; BMI, body mass index; TSC, time since smoking cessation.

