## Supplementary material for "Modelling the Complex Smoking Exposure History in Assessment of Pan-Cancer Risk": Table S and Figure S

**Author List and Affiliations**

Wei Liu [#], Ya-Ting Chen [#], Baiwenrui Tao, Ying Lv, Yan-Xi Zhang, Hui-Ying Ren, Yu-Ting Zhang, Yu-Ping Fan, Meng-Han Li, Shi-Yuan Wang, Bing-Wei Chen, Wen-Chao Li, Frits van Osch, Maurice P. Zeegers, Qi-Rong Qin [*], Anke Wesselius [*], Evan Yi-Wen Yu [*]

*Correspondence to:

**Evan Yi-Wen Yu, PhD**

Mail address: 87 Dingjiaqiao, Gulou District, Nanjing 210009, China.

ORCID: https://orcid.org/0000-0001-7825-5087

**&**

**Anke Wesselius, PhD**

Mailing address: Universiteitssingel 40 (Room C5.570), 6229 ER, Maastricht, the Netherlands.

ORCID: https://orcid.org/0000-0003-4474-9665

**&**

**Qi-Rong Qin, PhD**

Mail address: 849 JiangDong Road, Yushan District, Ma'anshan 243011, China.

### Content List

**Table S1.** Definitions of 12 cancers diagnosis in UK Biobank

**Table S2.** Serving size and coding of intake for each touchscreen food items/food groups

**Table S3.** Healthy diet score definition in UK Biobank

**Table S4.** AIC parameter and optimal model selection for 12 cancers

**Table S5.** General characteristics of the participants at baseline

**Table S6.** General characteristics of the participants at baseline by smoking

**Table S7.** Comparison of ERRs across 12 cancers when stratified for lifestyle behaviors

**Table S8.** The ratio of Min-Max range of ERRs/Max ERRs across 12 cancers

**Table S9.** Summary of published studies using flexible excess relative risk model to estimate complex smoking history and cancer risk

**Figure S1.** The excess relative risk (ERR) for pan-cancers per pack-year of smoking by smoking intensity (without adjustments)

**Figure S2.** The excess relative risk (ERR) for pan-cancers per pack-year of smoking by smoking intensity (adjusted for age, sex, BMI, ethnicity)

**Figure S3.** The excess relative risk (ERR) for pan-cancers per pack-year of smoking by smoking intensity (adjusted for age, sex, BMI, ethnicity, SES, alcohol consumption status, physical activity)

**Figure S4.** The excess relative risk (ERR) for pan-cancers per pack-year of smoking by smoking intensity (adjusted for age, sex, BMI, ethnicity, SES, alcohol consumption status, healthy diet)

**Figure S5.** The excess relative risk (ERR) for pan-cancers per pack-year of smoking by smoking intensity (adjusted for age, sex, BMI, ethnicity, SES, alcohol consumption status, physical activity, healthy diet)

**Figure S6.** Mean ERRs obtained based on main adjustment model across pan cancers

**Figure S7.** The excess relative risk (ERR) for pan-cancers per pack-year of smoking by time since smoking cessation (without adjustments)

**Figure S8.** The excess relative risk (ERR) for pan-cancers per pack-year of smoking by time since smoking cessation (adjusted for age, sex, BMI, ethnicity)

**Figure S9.** The excess relative risk (ERR) for pan-cancers per pack-year of smoking by time since smoking cessation (adjusted for age, sex, BMI, ethnicity, SES, alcohol consumption status, physical activity)

**Figure S10.** The excess relative risk (ERR) for pan-cancers per pack-year of smoking by time since smoking cessation (adjusted for age, sex, BMI, ethnicity, SES, alcohol consumption status, healthy diet)

**Figure S11.** The excess relative risk (ERR) for pan-cancers per pack-year of smoking by time since smoking cessation (adjusted for age, sex, BMI, ethnicity, SES, alcohol consumption status, physical activity, healthy diet)

**Figure S12.** The excess relative risk (ERR) for pan-cancers per pack-year of smoking by smoking intensity (limiting subjects to ages 50–74 years at enrollment and regrouping the former smokers who quitted smoking within 5 years at baseline as current smokers)

**Figure S13.** The excess relative risk (ERR) for pan-cancers per pack-year of smoking by smoking intensity (excluding non-White individuals)

**Figure S14.** The excess relative risk (ERR) for pan-cancers per pack-year of smoking by smoking intensity (excluding incident cases of cancer occurring during the first year of follow-up)

**Figure S15.** The excess relative risk (ERR) for pan-cancers per pack-year of smoking by smoking intensity (excluding the never smokers who reported with passive smoking)

**Figure S16.** The excess relative risk (ERR) for pan-cancers per pack-year of smoking by smoking intensity (considering the influence of frequency and amount of current alcohol consumption)

**Figure S17.** The excess relative risk (ERR) for pan-cancers per pack-year of smoking by time since smoking cessation. (limiting subjects to ages 50–74 years at enrollment and regrouping the former smokers who quitted smoking within 5 years at baseline as current smokers)

**Figure S18.** The excess relative risk (ERR) for pan-cancers per pack-year of smoking by time since smoking cessation (excluding non-White individuals)

**Figure S19.** The excess relative risk (ERR) for pan-cancers per pack-year of smoking by time since smoking cessation (excluding incident cases of cancer occurring during the first year of follow-up)

**Figure S20.** The excess relative risk (ERR) for pan-cancers per pack-year of smoking by time since smoking cessation (excluding the never smokers who reported with passive smoking)

**Figure S21.** The excess relative risk (ERR) for pan-cancers per pack-year of smoking by time since smoking cessation (considering the influence of frequency and amount of current alcohol consumption)

**Table S1.** Definitions of 12 cancers diagnosis in UK Biobank

| Disease | ICD-9 | ICD-10 |
|---|---|---|
| **Mouth and Throat** | 1400, 1401, 1403–1406, 1408–1416, 1418–1422, 1428–1431, 1438–1441, 1448–1456, 1458–1473, 1478–1483, 1488–1491, 1498, 1499 | C000–C006, C008, C009, C01, C020–C024, C028–C031, C039–C041, C048–C052, C058–C062, C068, C069, C07, C080, C081, C088–C091, C098–C104, C108–C113, C118, C119, C12, C130–C132, C138–C140, C142, C148 |
| **Larynx** | 1610 –1613, 1618, 1619 | C320–C323, C328, C329 |
| **Esophagus** | 1500–1505, 1508, 1509, 2301 | C150–C155, C158, C159, D001 |
| **Lung, Bronchus and Trachea** | 1622–1624, 1625, 1628, 1629, 2312 | C340–C343, C348, C349, D022 |
| **Stomach** | 1510–1516, 1518, 1519, 2302 | C160, C162–C166, C168, C169, D002 |
| **Liver** | 1550–1552, 2308 | C220–C224, C227, C229, D01500 |
| **Kidney and Renal Pelvis** | 1890, 1891 | C64, C65, D09101, D09102 |
| **Pancreas** | 1570–1574, 1578, 1579 | C250–C254, C257–C259, D01701 |
| **Colon and Rectum** | 2303, 2304, 1530–1541 | C180–C189, C19, C20, D010, D011, D012 |
| **Urinary Bladder** | 1880–1889, 2337 | C670, C671, C672, C673, C674, C675, C676, C677, C678, C679, D090 |
| **Uterine Cervix** | 1800, 1801, 1808, 1809, 2331 | C530, C531, C538, C539, D060, D061, D067, D069 |
| **Acute Myeloid Leukemia** | 2050 | C920 |

**Table S2.** Serving size and coding of intake for each touchscreen food items/food groups

| Food groups | Food items | Field IDs | Amount per serving | Coding |
|---|---|---|---|---|
| Red meat | Beef | 1369 (beef/week) | Once/week | 'Never' = 0, 'Less than once a week' = 0.5, 'Once a week' = 1, '2-4 times a week' = 3, '5-6 times a week' = 5.5, 'Once or more daily' = 7 |
| | Lamb/mutton | 1379 (lamb or mutton/week) | Once/week | 'Never' = 0, 'Less than once a week' = 0.5, 'Once a week' = 1, '2-4 times a week' = 3, '5-6 times a week' = 5.5, 'Once or more daily' = 7 |
| | Pork | 1389 (pork/week) | Once/week | 'Never' = 0, 'Less than once a week' = 0.5, 'Once a week' = 1, '2-4 times a week' = 3, '5-6 times a week' = 5.5, 'Once or more daily' = 7 |
| Processed meat | Processed meat | 1349 (processed meat/week or daily) | Once/week | 'Never' = 0, 'Less than once a week' = 0.5, 'Once a week' = 1, '2-4 times a week' = 3, '5-6 times a week' = 5.5, 'Once or more daily' = 7 |
| Total fish | Oily fish | 1329 (oily fish/week) | Once/week | 'Never' = 0, 'Less than once a week' = 0.5, 'Once a week' = 1, '2-4 times a week' = 3, '5-6 times a week' = 5.5, 'Once or more daily' = 7 |
| | Non-oily fish | 1339 (non-oily fish/week) | Once/week | 'Never' = 0, 'Less than once a week' = 0.5, 'Once a week' = 1, '2-4 times a week' = 3, '5-6 times a week' = 5.5, 'Once or more daily' = 7 |
| Total fruit | Fresh fruit | 1309 (pieces fresh fruit/day) | 1 piece | 'Less than one' = 0.5 |
| | Dried fruit | 1319 (pieces dried fruit/day) | 2 pieces | 'Less than one' = 0.5 |
| Total vegetables | Cooked vegetables | 1289 (tablespoons cooked vegetables/day) | 2 heaped tablespoons | 'Less than one' = 0.5 |
| | Salad/raw vegetables | 1299 (tablespoons salad/raw vegetables/day) | 2 heaped tablespoons | 'Less than one' = 0.5 |
| Whole grains | Whole meal/wholegrain bread | 1438-1 slice/week | 1 slice/day | 'Less than one' = 0.5 |
| | Bran/oat/muesli cereal | 1458-1 bowl/week | 1 bowl/day | 'Less than one' = 0.5 |
| Refined grains | White, brown, other bread | 1438-1 slice/week | 1 slice/day | 'Less than one' = 0.5 |
| | Biscuits, other cereals | 1458-1 bowl/week | 1 bowl/day | 'Less than one' = 0.5 |

**Table S3.** Healthy diet score definition in UK Biobank

| Healthy diet score factors | Intake goal | Field IDs | Amount per serving |
|---|---|---|---|
| Total fruits | ≥4 servings/day | 1309 (pieces fresh fruit/day)<br>1319 (pieces dried fruit/day) | 1309–1 piece<br>1319–2 pieces |
| Total vegetables | ≥4 servings/day | 1289 (tablespoons cooked vegetables/day)<br>1299 (salad/raw vegetables/day) | 2 heaped tablespoons |
| Total fish | ≥2 servings/week | 1329 (oily fish/week)<br>1339 (non-oily fish/week) | Once/week |
| Processed meats | ≤1 servings/week | 1349 (processed meat/week or daily) | Once/week |
| Red meat | ≤1.5 servings/week | 1369 (beef/week)<br>1379 (lamb or mutton/week)<br>1389 (pork/week) | Once/week |
| Whole grains | ≥3 servings/day | 1438, 1448 (whole meal/wholegrain bread slices/week)<br>1458, 1468 (bran/oat/muesli cereal bowls/week) | 1438/1448–1 slice/day,<br>1458/1468–1 bowl/day |
| Refined grains | ≤1.5 servings/day | 1438, 1448 (white, brown, other bread slices/week)<br>1458, 1468 (biscuits, other cereals/week) | 1438/1448–1 slice/day,<br>1458/1468–1 bowl/day |

**Table S4.** AIC parameter and optimal model selection for 12 cancers

| **Crude Model** | **G** | **AIC** | **β** | **μ1** | **μ2** | **μ3** | **μ4** |
|---|---|---|---|---|---|---|---|
| **Mouth and Throat** | G1 | 897.4 | | | | | |
| | **G2** | **893.9** | 0.966 | -1.754 | 0.363 | -1.045 | 0.170 |
| | G3 | 897.3 | | | | | |
| | G4 | 900.0 | | | | | |
| **Larynx** | G1 | 429.8 | | | | | |
| | G2 | 433.4 | | | | | |
| | G3 | 432.8 | | | | | |
| | **G4** | **429.0** | 1.008 | -0.050 | 0.001 | -0.090 | 0.002 |
| **Esophagus** | G1 | 1066.5 | | | | | |
| | **G2** | **1055.9** | 0.065 | -0.105 | 0.064 | -0.662 | 0.221 |
| | G3 | 1058.4 | | | | | |
| | G4 | 1061.8 | | | | | |
| **Lung, Bronchus and Trachea** | **G1** | **2118.6** | 0.679 | -0.141 | -0.327 | | |
| | G2 | 2119.5 | | | | | |
| | G3 | 2120.9 | | | | | |
| | G4 | 2131.7 | | | | | |
| **Stomach** | G1 | 897.5 | | | | | |
| | **G2** | **893.5** | 0.275 | -0.920 | 0.134 | -0.561 | 0.203 |
| | G3 | 894.4 | | | | | |
| | G4 | 896.1 | | | | | |
| **Liver** | G1 | 823.2 | | | | | |
| | G2 | 825.4 | | | | | |
| | G3 | 824.0 | | | | | |
| | **G4** | **821.4** | 0.008 | 0.087 | -0.001 | 0.012 | 0.001 |
| **Kidney and Renal Pelvis** | G1 | 1114.6 | | | | | |
| | **G2** | **1105.8** | 0.003 | 2.066 | -0.388 | -0.804 | 0.282 |
| | G3 | 1108.2 | | | | | |
| | G4 | 1108.7 | | | | | |
| **Pancreas** | G1 | 979.3 | | | | | |
| | G2 | 978.3 | | | | | |
| | G3 | 976.1 | | | | | |
| | **G4** | **971.9** | 0.028 | 0.032 | -0.001 | -0.075 | 0.003 |
| **Colon and Rectum** | G1 | 1969.1 | | | | | |
| | G2 | 1967.8 | | | | | |
| | G3 | 1961.1 | | | | | |
| | **G4** | **1960.9** | 0.024 | -0.038 | 0.000 | 0.048 | 0.000 |
| **Urinary Bladder** | G1 | 1609.7 | | | | | |
| | **G2** | **1562.4** | 0.196 | -0.571 | 0.062 | -0.764 | 0.299 |
| | G3 | 1573.0 | | | | | |
| | G4 | 1574.4 | | | | | |
| **Uterine Cervix** | **G1** | **539.6** | 0.012 | 0.376 | -29.793 | | |
| | G2 | 542.5 | | | | | |
| | G3 | 542.4 | | | | | |
| | G4 | 542.1 | | | | | |
| **Acute Myeloid Leukemia** | **G1** | **558.6** | 0.006 | 0.063 | 0.621 | | |
| | G2 | 561.9 | | | | | |
| | G3 | 560.5 | | | | | |
| | G4 | 559.5 | | | | | |
| **Model1** | | | | | | | |
| **Mouth and Throat** | **G1** | **3245.0** | 0.040 | 0.355 | -0.628 | | |
| | G2 | 3244.4 | | | | | |
| | G3 | 3246.1 | | | | | |
| | G4 | 3249.0 | | | | | |
| **Larynx** | G1 | 1179.8 | | | | | |
| | G2 | 1181.8 | | | | | |
| | G3 | 1180.4 | | | | | |
| | **G4** | **1177.7** | 1.043 | -0.075 | 0.001 | -0.069 | 0.000 |
| **Esophagus** | **G1** | **3240.3** | 0.029 | 0.222 | -0.236 | | |
| | G2 | 3242.1 | | | | | |
| | G3 | 3243.2 | | | | | |
| | G4 | 3245.6 | | | | | |
| **Lung, Bronchus and Trachea** | G1 | 8311.8 | | | | | |
| | G2 | 8305.6 | | | | | |
| | **G3** | **8298.6** | 0.821 | -0.294 | 0.011 | -0.263 | -0.030 |
| | G4 | 8311.3 | | | | | |
| **Stomach** | **G1** | **2709.5** | 0.119 | -0.337 | -0.245 | | |
| | G2 | 2712.8 | | | | | |
| | G3 | 2713.0 | | | | | |
| | G4 | 2714.6 | | | | | |

| | | | | | | | |
|---|---|---|---|---|---|---|---|
| **Liver** | **G1** | **2481.9** | 0.015 | 0.163 | -0.056 | | |
| | G2 | 2485.9 | | | | | |
| | G3 | 2486.3 | | | | | |
| | G4 | 2482.9 | | | | | |
| **Kidney and Renal Pelvis** | **G1** | **3695.7** | 0.065 | -0.283 | -0.478 | | |
| | G2 | 3697.6 | | | | | |
| | G3 | 3707.6 | | | | | |
| | G4 | 3701.1 | | | | | |
| **Pancreas** | **G1** | **3409.5** | 0.076 | -0.256 | -0.505 | | |
| | G2 | 3409.5 | | | | | |
| | G3 | 3409.1 | | | | | |
| | G4 | 3409.4 | | | | | |
| **Colon and Rectum** | G1 | 8519.1 | | | | | |
| | **G2** | **8518.8** | 0.025 | -0.857 | 0.131 | 0.756 | -0.184 |
| | G3 | 8520.3 | | | | | |
| | G4 | 8522.1 | | | | | |
| **Urinary Bladder** | **G1** | **5518.0** | 0.142 | -0.431 | -0.097 | | |
| | G2 | 5518.5 | | | | | |
| | G3 | 5523.4 | | | | | |
| | G4 | 5530.6 | | | | | |
| **Uterine Cervix** | **G1** | **1372.3** | 0.013 | 0.345 | -24.040 | | |
| **Acute Myeloid Leukemia** | **G1** | **1447.5** | 0.002 | 0.435 | 0.224 | | |
| | G2 | 1459.9 | | | | | |
| | G3 | 1460.1 | | | | | |
| | G4 | 1460.6 | | | | | |
| **Model2** | | | | | | | |
| **Mouth and Throat** | **G1** | **4997.0** | 0.037 | 0.340 | -0.634 | | |
| | G2 | 4997.0 | | | | | |
| | G3 | 4998.8 | | | | | |
| | G4 | 5001.9 | | | | | |
| **Larynx** | G1 | 1597.4 | | | | | |
| | G2 | 1599.2 | | | | | |
| | G3 | 1598.3 | | | | | |
| | **G4** | **1595.5** | 0.853 | -0.079 | 0.001 | -0.052 | 0.000 |
| **Esophagus** | **G1** | **5231.8** | 0.027 | 0.192 | -0.208 | | |
| | G2 | 5233.8 | | | | | |
| | G3 | 5234.9 | | | | | |
| | G4 | 5236.8 | | | | | |
| **Lung, Bronchus and Trachea** | G1 | 14519.0 | | | | | |
| | G2 | 14513.0 | | | | | |
| | **G3** | **14507.0** | 0.751 | -0.297 | 0.009 | -0.235 | -0.031 |
| | G4 | 14518.0 | | | | | |
| **Stomach** | **G1** | **4247.9** | 0.109 | -0.369 | -0.217 | | |
| | G2 | 4251.4 | | | | | |
| | G3 | 4252.3 | | | | | |
| | G4 | 4252.8 | | | | | |
| **Liver** | **G1** | **3796.8** | 0.012 | 0.152 | -0.013 | | |
| | G2 | 3800.8 | | | | | |
| | G3 | 3802.0 | | | | | |
| | G4 | 3797.7 | | | | | |
| **Kidney and Renal Pelvis** | **G1** | **5850.5** | 0.061 | -0.281 | -0.490 | | |
| | G2 | 5852.4 | | | | | |
| | G3 | 5854.5 | | | | | |
| | G4 | 5856.0 | | | | | |
| **Pancreas** | **G1** | **5278.8** | 0.070 | -0.270 | -0.507 | | |
| | G2 | 5282.8 | | | | | |
| | G3 | 5282.1 | | | | | |
| | G4 | 5282.4 | | | | | |
| **Colon and Rectum** | G1 | 14720.0 | | | | | |
| | **G2** | **14719.0** | 0.025 | -0.869 | 0.132 | 0.783 | -0.189 |
| | G3 | 14722.0 | | | | | |
| | G4 | 14722.0 | | | | | |
| **Urinary Bladder** | **G1** | **8950.0** | 0.134 | -0.432 | -0.090 | | |
| | G2 | 8950.6 | | | | | |
| | G3 | 8955.3 | | | | | |
| | G4 | 8962.4 | | | | | |
| **Uterine Cervix** | **G1** | **2263.4** | 0.014 | 0.215 | -1.383 | | |
| **Acute Myeloid Leukemia** | **G1** | **2152.3** | 0.001 | 0.546 | 0.245 | | |
| | G4 | 2154.9 | | | | | |
| **Model3** | | | | | | | |
| **Mouth and Throat** | **G1** | **6084.6** | 0.037 | 0.333 | -0.628 | | |

| | | | | | | | |
|---|---|---|---|---|---|---|---|
| | G2 | 6085.0 | | | | | |
| | G3 | 6086.4 | | | | | |
| | G4 | 6089.4 | | | | | |
| **Larynx** | G1 | 1873.0 | | | | | |
| | G2 | 1874.8 | | | | | |
| | G3 | 1874.0 | | | | | |
| | **G4** | **1871.5** | 0.820 | -0.079 | 0.001 | -0.048 | 0.000 |
| **Esophagus** | **G1** | **6378.0** | 0.027 | 0.188 | -0.205 | | |
| | G2 | 6380.3 | | | | | |
| | G3 | 6381.1 | | | | | |
| | G4 | 6382.8 | | | | | |
| **Lung, Bronchus and Trachea** | G1 | 18582.0 | | | | | |
| | G2 | 18576.0 | | | | | |
| | **G3** | **18570.0** | 0.742 | -0.296 | 0.009 | -0.228 | -0.031 |
| | G4 | 18581.0 | | | | | |
| **Stomach** | **G1** | **5229.5** | 0.108 | -0.367 | -0.218 | | |
| | G2 | 5233.0 | | | | | |
| | G3 | 5233.7 | | | | | |
| | G4 | 5234.5 | | | | | |
| **Liver** | **G1** | **4633.9** | 0.012 | 0.148 | -0.011 | | |
| | G2 | 4637.9 | | | | | |
| | G3 | 4639.0 | | | | | |
| | G4 | 4634.8 | | | | | |
| **Kidney and Renal Pelvis** | **G1** | **7329.4** | 0.061 | -0.288 | -0.485 | | |
| | G2 | 7331.3 | | | | | |
| | G3 | 7333.4 | | | | | |
| | G4 | 7334.8 | | | | | |
| **Pancreas** | **G1** | **6423.9** | 0.070 | -0.272 | -0.507 | | |
| | G2 | 6427.9 | | | | | |
| | G3 | 6427.1 | | | | | |
| | G4 | 6427.5 | | | | | |
| **Colon and Rectum** | G1 | 18929.0 | | | | | |
| | **G2** | **18928.0** | 0.025 | -0.886 | 0.135 | 0.780 | -0.187 |
| | G3 | 18931.0 | | | | | |
| | G4 | 18932.0 | | | | | |
| **Urinary Bladder** | **G1** | **11112.0** | 0.135 | -0.435 | -0.088 | | |
| | G2 | 11112.9 | | | | | |
| | G3 | 11117.0 | | | | | |
| | G4 | 11125.0 | | | | | |
| **Uterine Cervix** | **G1** | **2780.5** | 0.012 | 0.260 | -1.109 | | |
| **Acute Myeloid Leukemia** | **G1** | **2649.2** | 0.002 | 0.434 | 0.263 | | |
| | G2 | 2649.9 | | | | | |
| **Model 4** | | | | | | | |
| **Mouth and Throat** | **G1** | **6105.1** | 0.038 | 0.314 | -0.615 | | |
| | G2 | 6105.3 | | | | | |
| | G3 | 6107.0 | | | | | |
| | G4 | 6109.9 | | | | | |
| **Larynx** | G1 | 1916.3 | | | | | |
| | G2 | 1917.9 | | | | | |
| | G3 | 1917.1 | | | | | |
| | **G4** | **1914.3** | 0.790 | -0.084 | 0.001 | -0.036 | -0.001 |
| **Esophagus** | **G1** | **6485.5** | 0.026 | 0.180 | -0.188 | | |
| | G2 | 6487.7 | | | | | |
| | G3 | 6488.7 | | | | | |
| | G4 | 6490.1 | | | | | |
| **Lung, Bronchus and Trachea** | G1 | 18899.0 | | | | | |
| | G2 | 18892.0 | | | | | |
| | **G3** | **18886.0** | 0.754 | -0.316 | 0.009 | -0.215 | -0.032 |
| | G4 | 18896.0 | | | | | |
| **Stomach** | **G1** | **5268.9** | 0.107 | -0.379 | -0.204 | | |
| | G2 | 5272.4 | | | | | |
| | G3 | 5273.2 | | | | | |
| | G4 | 5273.9 | | | | | |
| **Liver** | **G1** | **4636.0** | 0.011 | 0.159 | 0.010 | | |
| | G2 | 4640.1 | | | | | |
| | G3 | 4641.3 | | | | | |
| | G4 | 4636.9 | | | | | |
| **Kidney and Renal Pelvis** | **G1** | **7361.0** | 0.058 | -0.269 | -0.494 | | |
| | G2 | 7363.1 | | | | | |
| | G3 | 7365.4 | | | | | |
| | G4 | 7366.6 | | | | | |

| | | | | | | | |
|---|---|---|---|---|---|---|---|
| **Pancreas** | **G1** | **6580.1** | 0.071 | -0.272 | -0.508 | | |
| | G2 | 6584.1 | | | | | |
| | G3 | 6583.4 | | | | | |
| | G4 | 6583.7 | | | | | |
| **Colon and Rectum** | G1 | 19386.0 | | | | | |
| | **G2** | **19385.0** | 0.023 | 0.995 | 0.152 | 0.967 | -0.226 |
| | G3 | 19387.0 | | | | | |
| | G4 | 19388.0 | | | | | |
| **Urinary Bladder** | **G1** | **11339.0** | 0.136 | -0.446 | -0.079 | | |
| | G2 | 11339.0 | | | | | |
| | G3 | 11344.0 | | | | | |
| | G4 | 11351.0 | | | | | |
| **Uterine Cervix** | **G1** | **2888.4** | 0.010 | 0.365 | -23.687 | | |
| **Acute Myeloid Leukemia** | **G1** | **2668.7** | 0.002 | 0.494 | 0.233 | | |
| | G2 | 2669.2 | | | | | |
| **Model5** | | | | | | | |
| **Mouth and Throat** | **G1** | **7202.7** | 0.038 | 0.308 | -0.611 | | |
| | G2 | 7203.0 | | | | | |
| | G3 | 7204.7 | | | | | |
| | G4 | 7207.5 | | | | | |
| **Larynx** | G1 | 2173.0 | | | | | |
| | G2 | 2174.6 | | | | | |
| | G3 | 2173.5 | | | | | |
| | **G4** | **2171.2** | 0.773 | -0.084 | 0.001 | -0.034 | -0.001 |
| **Esophagus** | **G1** | **7667.0** | 0.026 | 0.177 | -0.185 | | |
| | G2 | 7669.2 | | | | | |
| | G3 | 7670.1 | | | | | |
| | G4 | 7671.6 | | | | | |
| **Lung, Bronchus and Trachea** | G1 | 22952.0 | | | | | |
| | G2 | 22945.0 | | | | | |
| | **G3** | **22939.0** | 0.761 | -0.323 | 0.009 | -0.210 | -0.032 |
| | G4 | 22949.0 | | | | | |
| **Stomach** | **G1** | **6265.0** | 0.104 | -0.371 | -0.206 | | |
| | G2 | 6268.5 | | | | | |
| | G3 | 6269.2 | | | | | |
| | G4 | 6269.9 | | | | | |
| **Liver** | **G1** | **5464.5** | 0.011 | 0.156 | 0.012 | | |
| | G2 | 5468.5 | | | | | |
| | G3 | 5469.7 | | | | | |
| | G4 | 5465.1 | | | | | |
| **Kidney and Renal Pelvis** | **G1** | **8966.5** | 0.059 | -0.276 | -0.487 | | |
| | G2 | 8968.8 | | | | | |
| | G3 | 8970.5 | | | | | |
| | G4 | 8972.0 | | | | | |
| **Pancreas** | **G1** | **7790.8** | 0.072 | -0.276 | -0.509 | | |
| | G2 | 7794.8 | | | | | |
| | G3 | 7794.1 | | | | | |
| | G4 | 7794.5 | | | | | |
| **Colon and Rectum** | G1 | 23921.0 | | | | | |
| | **G2** | **23920.0** | 0.023 | -0.998 | 0.152 | 0.967 | -0.224 |
| | G3 | 23922.0 | | | | | |
| | G4 | 23924.0 | | | | | |
| **Urinary Bladder** | **G1** | **13600.0** | 0.139 | -0.455 | -0.077 | | |
| | G2 | 13600.6 | | | | | |
| | G3 | 13605.0 | | | | | |
| | G4 | 13611.0 | | | | | |
| **Uterine Cervix** | **G1** | **3455.9** | 0.010 | 0.363 | -17.153 | | |
| **Acute Myeloid Leukemia** | **G1** | **3159.9** | 0.002 | 0.436 | 0.246 | | |
| | G2 | 3160.4 | | | | | |

$G_1=\exp\{\mu_1\ln(n)+\mu_2\ln(t)\}$

$G_2=\exp\{\mu_1\ln(n)+\mu_2\ln(n)^2+\mu_3\ln(t)+\mu_4\ln(t)^2\}$

$G_3=\exp\{\mu_1\ln(n)+\mu_2 n+\mu_3\ln(t)+\mu_4 t\}$

$G_4=\exp\{\mu_1 n+\mu_2 n^2+\mu_3 t+\mu_4 t^2\}$

**Abbreviations**: AIC, Akaike information criterion.

**Table S5.** General characteristics of the participants at baseline

| Characteristic | Total n (%) | Non-cancer control n (%) | Case n (%) |
|---|---|---|---|
| **Median follow up years** | | 11.63 | 4.93 |
| **Age (years), mean ± SD** | 56.99 ± 8.09 | 56.85 ± 8.10 | 61.34 ± 6.64 |
| < 45 | 50,746 (10.33) | 50,331 (10.58) | 415 (2.72) |
| 45–50 | 65,257 (13.29) | 64,496 (13.55) | 761 (4.99) |
| 50–55 | 75,097 (15.29) | 73,631 (15.47) | 1,466 (9.61) |
| 55–60 | 88,815 (18.09) | 86,298 (18.14) | 2,517 (16.5) |
| 60–65 | 118,028 (24.04) | 113,381 (23.83) | 4,647 (30.47) |
| ≥65 | 93,119 (18.96) | 87,673 (18.43) | 5,446 (35.71) |
| **Sex** | | | |
| Women | 270,147 (55.01) | 263,687 (55.42) | 6,460 (42.36) |
| Men | 220,915 (44.99) | 212,123 (44.58) | 8,792 (57.64) |
| **BMI (kg/m$^2$)** | | | |
| < 18.5 | 2,583 (0.53) | 2,481 (0.52) | 102 (0.67) |
| 18.5–25 | 159,945 (32.57) | 155,759 (32.74) | 4,186 (27.45) |
| 25–30 | 207,121 (42.18) | 200,527 (42.14) | 6,594 (43.23) |
| ≥30 | 118,881 (24.21) | 114,611 (24.09) | 4,270 (28.00) |
| unknown | 2,532 (0.52) | 2,432 (0.51) | 100 (0.65) |
| **Ethnicity** * | | | |
| Ethnicity 1 | 462,873 (94.26) | 448,180 (94.19) | 14,693 (96.33) |
| Ethnicity 2 | 2,913 (0.59) | 2,837 (0.60) | 76 (0.50) |
| Ethnicity 3 | 9,678 (1.97) | 9,509 (2.00) | 169 (1.11) |
| Ethnicity 4 | 7,912 (1.61) | 7,786 (1.64) | 126 (0.83) |
| Ethnicity 5 | 1,559 (0.32) | 1,526 (0.32) | 33 (0.22) |
| Ethnicity 6 | 4,460 (0.91) | 4,359 (0.92) | 101 (0.66) |
| Ethnicity 7 | 1,667 (0.34) | 1,613 (0.34) | 54 (0.35) |
| **Socioeconomic status** | | | |
| high | 121,985 (24.84) | 119,326 (25.08) | 2,659 (17.43) |
| median | 119,638 (24.36) | 116,580 (24.50) | 3,058 (20.05) |
| low | 113,619 (23.14) | 109,896 (23.10) | 3,723 (24.41) |
| unknown | 135,820 (27.66) | 130,008 (27.32) | 5,812 (38.11) |
| **Alcohol consumption status** | | | |
| never | 22,086 (4.50) | 21,507 (4.52) | 579 (3.80) |
| previous | 17,619 (3.59) | 16,779 (3.53) | 840 (5.51) |
| current | 450,814 (91.80) | 436,996 (91.84) | 13,818 (90.60) |
| unknown | 543 (0.11) | 528 (0.11) | 15 (0.10) |
| **Alcohol consumption** | | | |
| ≤1 drink/day for women<br>≤2drinks/day for men | 283,371 (57.71) | 275,079 (57.82) | 8,292 (54.37) |
| >1drink/day for women<br>>2drinks/day for men | 139,040 (28.31) | 134,382 (28.24) | 4,658 (30.54) |
| unknown | 68,651 (13.98) | 66,349 (13.94) | 2,302 (15.09) |
| **Diet** | | | |
| unhealthy diet | 213,508 (43.48) | 206,597 (43.42) | 6,911 (45.31) |
| healthy diet | 213,168 (43.41) | 207,452 (43.60) | 5,716 (37.48) |
| unknown | 64,386 (13.11) | 61,761 (12.98) | 2,625 (17.21) |
| **Physical activity** | | | |
| un-regular physical activity | 73,213 (14.91) | 70,776 (14.87) | 2,437 (15.98) |
| regular physical activity | 321,885 (65.55) | 312,308 (65.64) | 9,577 (62.79) |
| unknown | 95,964 (19.54) | 92,726 (19.49) | 3,238 (21.23) |

* Ethnicity 1: “White”, “British”, “Irish”, “Any other white background”; Ethnicity 2: “Mixed”, “White and Black Caribbean”, “White and Black African”, “White and Asian”, “Any other mixed background”; ethnicity 3: “Asian or Asian British”, “Indian”, “Pakistani”, “Bangladeshi”, “Any other Asian background”; Ethnicity 4: “Black or Black British”, “Caribbean”, “African”, “Any other Black background”; Ethnicity 5: “Chinese”; Ethnicity 6: “Other ethnic group”; Ethnicity 7: “Prefer not to answer”, “Do not know”.

**Table S6.** General characteristics of the participants at baseline by smoking

| Characteristic | Never | Current | Former | Current | | Former | | |
|---|---|---|---|---|---|---|---|---|
| | | | | Intensity>10 | Duration>20 | Intensity>10 | Duration>20 | TSC>5 |
| **Cancer cases** | 7,519 (2.22) | 2,504 (6.69) | 5,229 (4.57) | 1,878 (75.00) | 2,410 (96.25) | 4,304 (82.31) | 3,499 (66.92) | 4,283 (81.91) |
| **Non-cancer controls** | 331,680 (97.78) | 34,937 (93.31) | 109,193 (95.43) | 22,363 (64.01) | 32,780 (93.83) | 80,267 (73.51) | 55,039 (50.41) | 92,874 (85.05) |
| **Age (years), mean ± SD** | 56.57 ± 8.15 | 55.17 ± 8.06 | 58.81 ± 7.62 | 55.39 ± 7.92 | 55.36 ± 7.98 | 59.05 ± 7.42 | 60.00 ± 7.18 | 59.43 ± 7.32 |
| < 45 | 37,704 (11.12) | 5,149 (13.75) | 7,893 (6.90) | 3,036 (12.52) | 4,494 (12.77) | 5,028 (5.95) | 2,496 (4.26) | 5,215 (5.37) |
| 45–50 | 48,452 (14.28) | 6,302 (16.83) | 10,503 (9.18) | 3,980 (16.42) | 5,877 (16.70) | 7,353 (8.69) | 4,564 (7.80) | 7,858 (8.09) |
| 50–55 | 53,915 (15.89) | 6,675 (17.83) | 14,507 (12.68) | 4,419 (18.23) | 6,351 (18.05) | 10,831 (12.81) | 6,738 (11.51) | 11,709 (12.05) |
| 55–60 | 60,776 (17.92) | 6,816 (18.20) | 21,223 (18.55) | 4,573 (18.86) | 6,516 (18.52) | 16,093 (19.03) | 10,116 (17.28) | 18,237 (18.77) |
| 60–65 | 77,747 (22.92) | 7,562 (20.20) | 32,719 (28.60) | 5,088 (20.99) | 7,232 (20.55) | 24,645 (29.14) | 17,636 (30.13) | 28,918 (29.76) |
| ≥65 | 60,605 (17.87) | 4,937 (13.19) | 27,577 (24.10) | 3,145 (12.97) | 4,720 (13.41) | 20,621 (24.38) | 16,988 (29.02) | 25,220 (25.96) |
| **Sex** | | | | | | | | |
| Women | 197,581 (58.25) | 18,277 (48.82) | 54,289 (47.45) | 10,742 (44.31) | 17,515 (49.77) | 36,494 (43.15) | 26,899 (45.95) | 45,598 (46.93) |
| Men | 141,618 (41.75) | 19,164 (51.18) | 60,133 (52.55) | 13,499 (55.69) | 17,675 (50.23) | 48,077 (56.85) | 31,639 (54.05) | 51,559 (53.07) |
| **BMI (kg/m$^2$)** | | | | | | | | |
| < 18.5 | 1,753 (0.52) | 510 (1.36) | 320 (0.28) | 326 (1.34) | 498 (1.42) | 198 (0.23) | 158 (0.27) | 251 (0.26) |
| 18.5-25 | 117,465 (34.63) | 13,650 (36.46) | 28,830 (25.20) | 8,404 (34.67) | 12,887 (36.62) | 18,416 (21.78) | 12,073 (20.62) | 24,586 (25.31) |
| 25 - 30 | 141,421 (41.69) | 14,807 (39.55) | 50,893 (44.48) | 9,601 (39.61) | 13,856 (39.37) | 37,821 (44.72) | 26,177 (44.72) | 43,356 (44.62) |
| ≥30 | 76,884 (22.67) | 8,131 (21.72) | 33,866 (29.60) | 5,678 (23.42) | 7,623 (21.66) | 27,746 (32.81) | 19,849 (33.91) | 28,557 (29.39) |
| unknown | 1,676 (0.49) | 343 (0.92) | 513 (0.45) | 232 (0.96) | 326 (0.93) | 390 (0.46) | 281 (0.48) | 407 (0.42) |
| **Ethnicity** * | | | | | | | | |
| Ethnicity1* | 316,528 (93.32) | 35,241 (94.12) | 111,104 (97.10) | 23,271 (96.00) | 33,183 (94.30) | 82,553 (97.61) | 56,803 (97.04) | 94,632 (97.40) |
| Ethnicity2* | 1,859 (0.55) | 426 (1.14) | 628 (0.55) | 219 (0.90) | 391 (1.11) | 420 (0.50) | 342 (0.58) | 468 (0.48) |
| Ethnicity3* | 8,356 (2.46) | 547 (1.46) | 775 (0.68) | 235 (0.97) | 485 (1.38) | 465 (0.55) | 413 (0.71) | 586 (0.6) |
| Ethnicity4* | 6,592 (1.94) | 604 (1.61) | 716 (0.63) | 188 (0.78) | 558 (1.59) | 321 (0.38) | 377 (0.64) | 515 (0.53) |
| Ethnicity5* | 1,364 (0.40) | 88 (0.24) | 107 (0.09) | 37 (0.15) | 84 (0.24) | 58 (0.07) | 46 (0.08) | 88 (0.09) |
| Ethnicity6* | 3,366 (0.99) | 376 (1.00) | 718 (0.63) | 193 (0.80) | 343 (0.97) | 473 (0.56) | 361 (0.62) | 558 (0.57) |
| Ethnicity7* | 1,134 (0.33) | 159 (0.42) | 374 (0.33) | 98 (0.40) | 146 (0.41) | 281 (0.33) | 196 (0.33) | 310 (0.32) |
| **Socioeconomic status** | | | | | | | | |
| high | 94,708 (27.92) | 3,953 (10.56) | 23,324 (20.38) | 2,135 (8.81) | 3,525 (10.02) | 16,226 (19.19) | 8,427 (14.40) | 20,855 (21.47) |
| median | 85,011 (25.06) | 7,020 (18.75) | 27,607 (24.13) | 4,118 (16.99) | 6,421 (18.25) | 19,677 (23.27) | 12,538 (21.42) | 23,757 (24.45) |
| low | 71,932 (21.21) | 12,866 (34.36) | 28,821 (25.19) | 8,583 (35.41) | 12,158 (34.55) | 21,993 (26.01) | 16,109 (27.52) | 23,382 (24.07) |
| unknown | 87,548 (25.81) | 13,602 (36.33) | 34,670 (30.30) | 9,405 (38.80) | 13,086 (37.19) | 26,675 (31.54) | 21,464 (36.67) | 29,163 (30.02) |
| **Alcohol consumption status** | | | | | | | | |
| never | 19,247 (5.67) | 1,118 (2.99) | 1,721 (1.50) | 698 (2.88) | 1,050 (2.98) | 1,262 (1.49) | 1,106 (1.89) | 1,331 (1.37) |
| previous | 9,992 (2.95) | 2,384 (6.37) | 5,243 (4.58) | 1,732 (7.14) | 2,295 (6.52) | 4,199 (4.97) | 3,058 (5.22) | 4,271 (4.40) |
| current | 309,592 (91.27) | 33,835 (90.37) | 107,387 (93.85) | 21,736 (89.67) | 31,750 (90.22) | 79,055 (93.48) | 54,330 (92.81) | 91,502 (94.18) |
| prefer not to | 368 | 104 | 71 | 75 | 95 | 55 | 44 | 53 |

| | | | | | | | | |
|---|---|---|---|---|---|---|---|---|
| answer | (0.11) | (0.28) | (0.06) | (0.31) | (0.27) | (0.07) | (0.08) | (0.05) |
| **Alcohol consumption** | | | | | | | | |
| ≤1 drink/day for women ≤2drinks/day for men | 209,156 (61.66) | 16,824 (44.93) | 57,391 (50.16) | 10,573 (43.62) | 15,764 (44.80) | 41,773 (49.39) | 28,439 (48.58) | 49,390 (50.84) |
| >1drink/day for women >2drinks/day for men | 81,405 (24.00) | 13,827 (36.93) | 43,808 (38.29) | 9,083 (37.47) | 12,929 (36.74) | 32,847 (38.84) | 22,307 (38.11) | 37,363 (38.46) |
| unknown | 48,638 (14.34) | 6,790 (18.14) | 13,223 (11.56) | 4,585 (18.91) | 6,497 (18.46) | 9,951 (11.77) | 7,792 (13.31) | 10,404 (10.71) |
| **Diet** | | | | | | | | |
| unhealthy diet | 146,257 (43.12) | 16,880 (45.08) | 50,371 (44.02) | 11,152 (46.00) | 15,861 (45.07) | 38,162 (45.12) | 26,633 (45.50) | 42,606 (43.86) |
| healthy diet | 155,325 (45.79) | 9,484 (25.33) | 48,359 (42.26) | 5,008 (20.66) | 8,810 (25.04) | 33,965 (40.16) | 22,669 (38.72) | 42,227 (43.46) |
| unknown | 37,617 (11.09) | 11,077 (29.59) | 15,692 (13.71) | 8,081 (33.34) | 10,519 (29.89) | 12,444 (14.72) | 9,236 (15.78) | 12,324 (12.68) |
| **Physical activity** | | | | | | | | |
| un-regular physical activity | 49,223 (14.51) | 6,558 (17.52) | 17,432 (15.23) | 4,418 (18.23) | 6,136 (17.44) | 13,384 (15.83) | 9,106 (15.56) | 14,450 (14.87) |
| regular physical activity | 223,909 (66.01) | 22,284 (59.52) | 75,692 (66.15) | 14,021 (57.84) | 20,864 (59.29) | 55,390 (65.50) | 37,721 (64.44) | 64,769 (66.66) |
| unknown | 66,067 (19.48) | 8,599 (22.97) | 21,298 (18.61) | 5,802 (23.93) | 8,190 (23.27) | 15,797 (18.68) | 11,711 (20.01) | 17,938 (18.46) |

* Ethnicity 1: "White", "British", "Irish", "Any other white background"; Ethnicity 2: "Mixed", "White and Black Caribbean", "White and Black African", "White and Asian", "Any other mixed background"; ethnicity 3: "Asian or Asian British", "Indian", "Pakistani", "Bangladeshi", "Any other Asian background"; Ethnicity 4: "Black or Black British", "Caribbean", "African", "Any other Black background"; Ethnicity 5: "Chinese"; Ethnicity 6: "Other ethnic group"; Ethnicity 7: "Prefer not to answer", "Do not know".
**Abbreviations:** TSC, time since smoking cessation.

**Table S7.** Comparison of ERRs across 12 cancers when stratified for lifestyle behaviors

| Cancer type | group | β | μ1 | μ2 | μ3 | μ4 | $p^*$ |
|---|---|---|---|---|---|---|---|
| Mouth and Throat | regular physical activity | 0.022 | 0.470 | -0.562 | | | <0.001 |
| | un-regular physical activity | 0.057 | 0.174 | -0.696 | | | |
| Larynx | regular physical activity | 1.253 | -0.099 | 0.001 | -0.074 | 0.001 | 0.003532 |
| | un-regular physical activity | 1.447 | -0.135 | 0.002 | 0.148 | -0.011 | |
| Esophagus | regular physical activity | 0.032 | 0.186 | -0.235 | | | <0.001 |
| | un-regular physical activity | 0.102 | -0.355 | -0.235 | | | |
| Lung, Bronchus and Trachea | regular physical activity | 0.486 | -0.102 | -0.001 | -0.167 | -0.042 | <0.001 |
| | un-regular physical activity | 3.298 | -1.094 | 0.047 | -0.015 | -0.062 | |
| Stomach | regular physical activity | 0.090 | -0.256 | -0.299 | | | <0.001 |
| | un-regular physical activity | 0.136 | -0.549 | -0.061 | | | |
| Liver | regular physical activity | 0.020 | 0.028 | -0.038 | | | <0.001 |
| | un-regular physical activity | 0.003 | 0.560 | -0.145 | | | |
| Kidney and Renal Pelvis | regular physical activity | 0.076 | -0.395 | -0.447 | | | <0.001 |
| | un-regular physical activity | 0.000 | 1.022 | -0.158 | | | |
| Pancreas | regular physical activity | 0.047 | -0.090 | -0.592 | | | <0.001 |
| | un-regular physical activity | 0.001 | 1.283 | -1.283 | | | |
| Colon and Rectum | regular physical activity | 0.065 | -0.146 | 0.047 | -0.839 | 0.196 | <0.001 |
| | un-regular physical activity | 0.073 | -0.125 | -0.036 | -0.350 | 0.042 | |
| Urinary Bladder | regular physical activity | 0.123 | -0.369 | -0.146 | | | <0.001 |
| | un-regular physical activity | 0.401 | -0.797 | -0.008 | | | |
| Uterine Cervix | regular physical activity | 0.009 | 0.539 | -28.282 | | | <0.001 |
| | un-regular physical activity | 0.061 | -1.014 | 0.061 | | | |
| Acute Myeloid Leukemia | regular physical activity | 0.011 | 0.220 | -0.023 | | | 0.005801 |
| | un-regular physical activity | 0.000 | -0.163 | 5.388 | | | |
| Mouth and Throat | healthy diet | 0.070 | 0.051 | -0.491 | | | <0.001 |
| | unhealthy diet | 0.149 | -0.180 | -0.463 | | | |
| Larynx | healthy diet | 0.347 | 0.124 | -0.010 | 0.113 | -0.008 | 0.04677 |
| | unhealthy diet | 0.634 | -0.104 | 0.002 | 0.121 | -0.011 | |
| Esophagus | healthy diet | 0.006 | 0.708 | -0.367 | | | <0.001 |
| | unhealthy diet | 0.035 | 0.121 | -0.228 | | | |
| Lung, Bronchus and Trachea | healthy diet | 0.921 | -0.429 | 0.015 | -0.346 | -0.013 | 0.5746 |
| | unhealthy diet | 0.971 | -0.405 | 0.015 | -0.227 | -0.033 | |
| Stomach | healthy diet | 0.048 | -0.226 | 0.007 | | | <0.001 |
| | unhealthy diet | 0.063 | -0.140 | -0.326 | | | |
| Liver | healthy diet | 0.008 | 0.495 | -0.446 | | | <0.001 |
| | unhealthy diet | 0.016 | 0.079 | 0.028 | | | |
| Kidney and Renal Pelvis | healthy diet | 0.037 | -0.151 | -0.347 | | | <0.001 |
| | unhealthy diet | 0.077 | -0.342 | -0.526 | | | |
| Pancreas | healthy diet | 0.088 | -0.341 | -0.651 | | | <0.001 |
| | unhealthy diet | 0.285 | -0.775 | -0.555 | | | |
| Colon and Rectum | healthy diet | 0.032 | -0.725 | 0.080 | 0.141 | 0.026 | 0.642 |
| | unhealthy diet | 0.016 | -1.174 | 0.190 | 1.091 | -0.210 | |
| Urinary Bladder | healthy diet | 0.318 | -0.773 | -0.074 | | | <0.001 |
| | unhealthy diet | 0.111 | -0.332 | -0.124 | | | |
| Uterine Cervix | healthy diet | 0.051 | 0.097 | -24.017 | | | 0.2963 |
| | unhealthy diet | 0.020 | -0.023 | -8.734 | | | |
| Acute Myeloid Leukemia | healthy diet | 0.002 | 0.854 | 0.030 | | | <0.001 |
| | unhealthy diet | 0.000 | -1.177 | 4.775 | | | |

$p^*$: Perform a t-test on the ERR/pack-year values for each two groups.

**Table S8.** The ratio of Min-Max range of ERRs/Max ERRs across 12 cancers

| Cancer type | TSC=10 years | TSC=20 years | TSC=30 years | TSC=40 years |
|---|---|---|---|---|
| Mouth and Throat | 63.97% | 76.78% | 82.05% | 85.04% |
| Larynx | 35.73% | 64.96% | 82.01% | 91.30% |
| Esophagus | 28.44% | 38.04% | 43.05% | 46.36% |
| Lung, Bronchus and Trachea | 46.41% | 66.50% | 77.59% | 84.59% |
| Stomach | 29.46% | 39.30% | 44.41% | 47.77% |
| Liver | 2.06% | 2.94% | 3.45% | 3.81% |
| Kidney and Renal Pelvis | 54.56% | 67.65% | 73.48% | 76.97% |
| Pancreas | 55.74% | 68.85% | 74.63% | 78.07% |
| Colon and Rectum | -41.37% | -21.29% | -1.92% | 13.25% |
| Urinary Bladder | 13.42% | 18.62% | 21.52% | 23.52% |
| Uterine Cervix | 89.20% | 95.86% | 97.64% | 98.41% |

**Abbreviations:** TSC, time since smoking cessation.

**Table S9.** Summary of published studies using flexible excess relative risk model to estimate complex smoking history and cancer risk

| Cancer type | Conclusion |
|---|---|
| Oral Cavity and Pharyngeal Cancer | The pack-years association (i.e., the EOR/pack-year estimates) diminished with increasing delivery rate above 15 cigarettes/day—that is, for fixed pack-years, a higher cigarettes/day for a shorter duration was less deleterious than a lower cigarettes/day for a longer duration [1]. |
| Laryngeal Cancer | In lowest BMI category, for fixed pack-years, a higher cigarettes/day for a shorter duration was less deleterious than a lower cigarettes/day for a longer duration [1]. |
| Lung Cancer | The EOR/pack-year increases with intensity for subjects who smoke ≤20 cigarettes per day and decreases with intensity for subjects who smoke >20 cigarettes per day [2];<br>For an equal total exposure (in pack-years), smoking at a lower intensity for a longer duration is more harmful than smoking at a higher intensity for a shorter duration [3]. |
| Bladder Cancer | For an equal total exposure (in pack-years), smoking at a lower intensity for a longer duration is more harmful than smoking at a higher intensity for a shorter duration [3,4]. |
| Pancreatic Cancer | For an equal total exposure (in pack-years), smoking at a lower intensity for a longer duration is more harmful than smoking at a higher intensity for a shorter duration [3,5,6]. |
| Esophageal Cancer | Excess RR/pack-year estimates increased with intensity at low intensities for the esophagus, and kidney but not for the other sites. The 95 percent confidence intervals suggested low power for estimating intensity effects below 10 cigarettes per day, because of few cases [3]. |
| Kidney Cancer | Excess RR/pack-year estimates increased with intensity at low intensities for the esophagus, and kidney but not for the other sites. The 95 percent confidence intervals suggested low power for estimating intensity effects below 10 cigarettes per day, because of few cases [3]. |
| Liver Cancer | For an equal total exposure (in pack-years), smoking at a lower intensity for a longer duration is more harmful than smoking at a higher intensity for a shorter duration [3]. |

**Figure S1.** The excess relative risk (ERR) for pan-cancers per pack-year of smoking by smoking intensity (without adjustments)

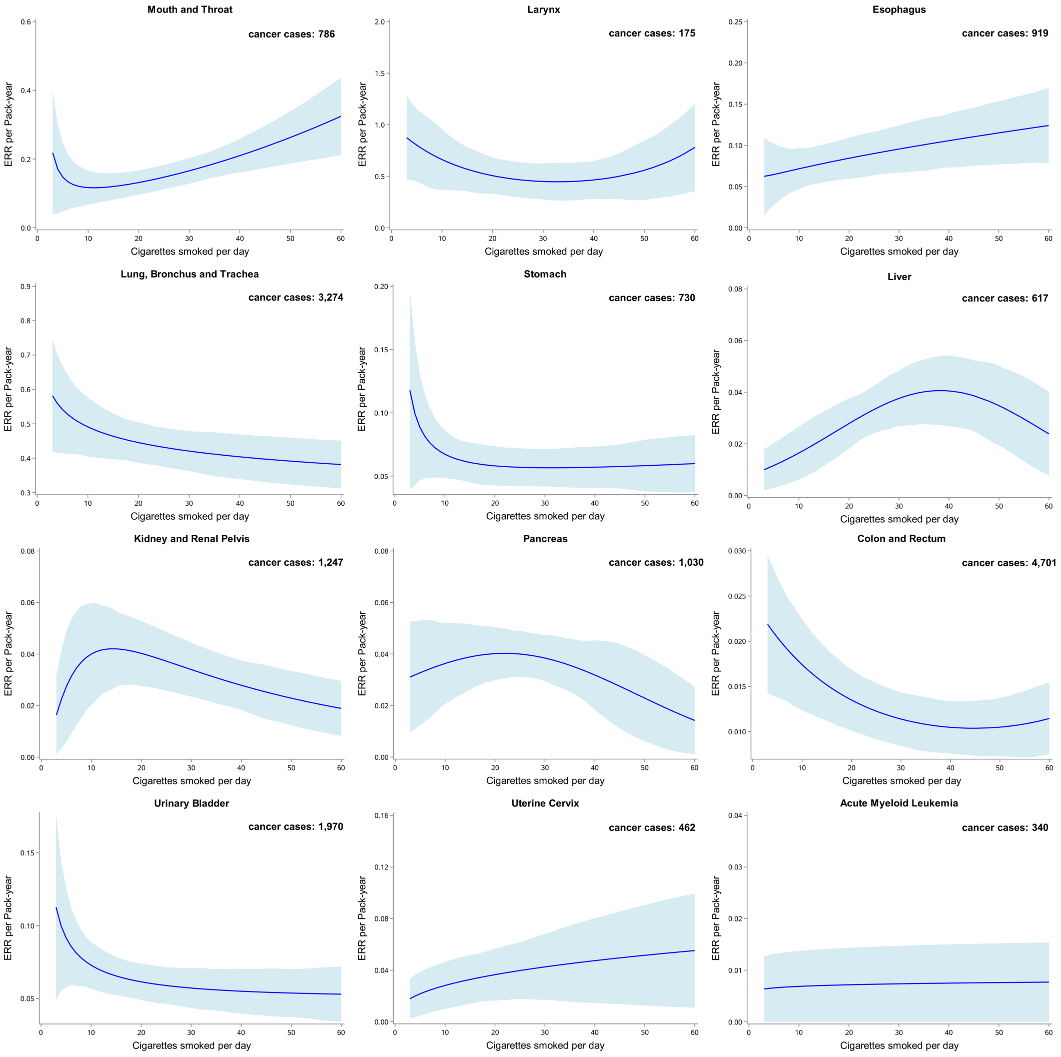

Bootstrapped 95% confidence intervals are based on 1,000 replications. The blue solid line represents the prediction from the function $g_1(n)$ in the model, with the area showing 95% confidence interval obtained from bootstrap analysis.
**Model adjustments:** without adjustments.
**Abbreviations:** ERR, excess relative risk.

**Figure S2.** The excess relative risk (ERR) for pan-cancers per pack-year of smoking by smoking intensity (adjusted for age, sex, BMI, ethnicity)

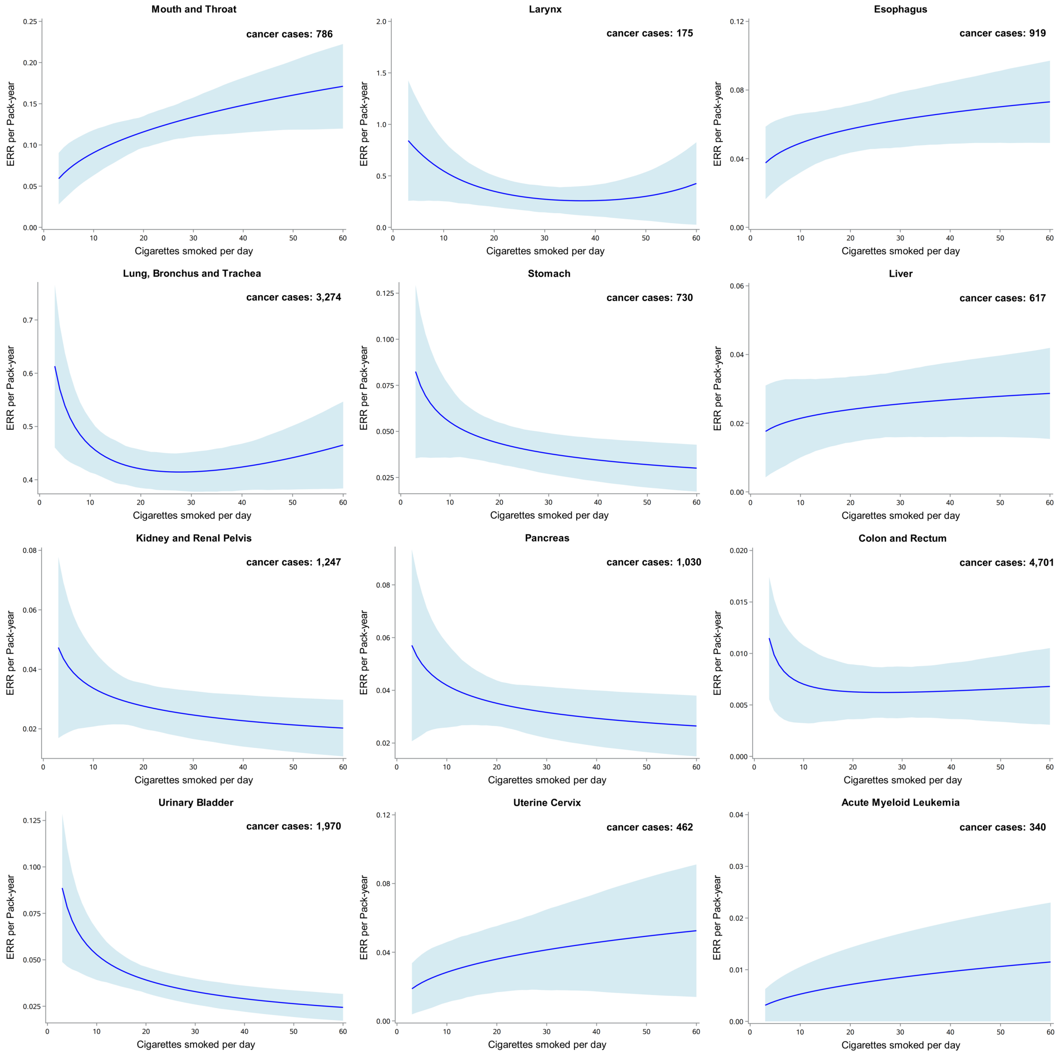

Bootstrapped 95% confidence intervals are based on 1,000 replications. The blue solid line represents the prediction from the function $g_1(n)$ in the model, with the area showing 95% confidence interval obtained from bootstrap analysis.

**Model adjustments:** age (<45, 45–50, 50–60, 60–65, ≥65, years), sex (men or women), ethnicity (White, Asian or Asian British, Black or Black British, Chinese, Mixed, other ethnic group, or unknown), BMI (<18.5, 18.5–25, 25–30, ≥30, or unknown, kg/m$^2$).

**Abbreviations:** ERR, excess relative risk; BMI, body mass index.

**Figure S3.** The excess relative risk (ERR) for pan-cancers per pack-year of smoking by smoking intensity (adjusted for age, sex, BMI, ethnicity, SES, alcohol consumption status, physical activity)

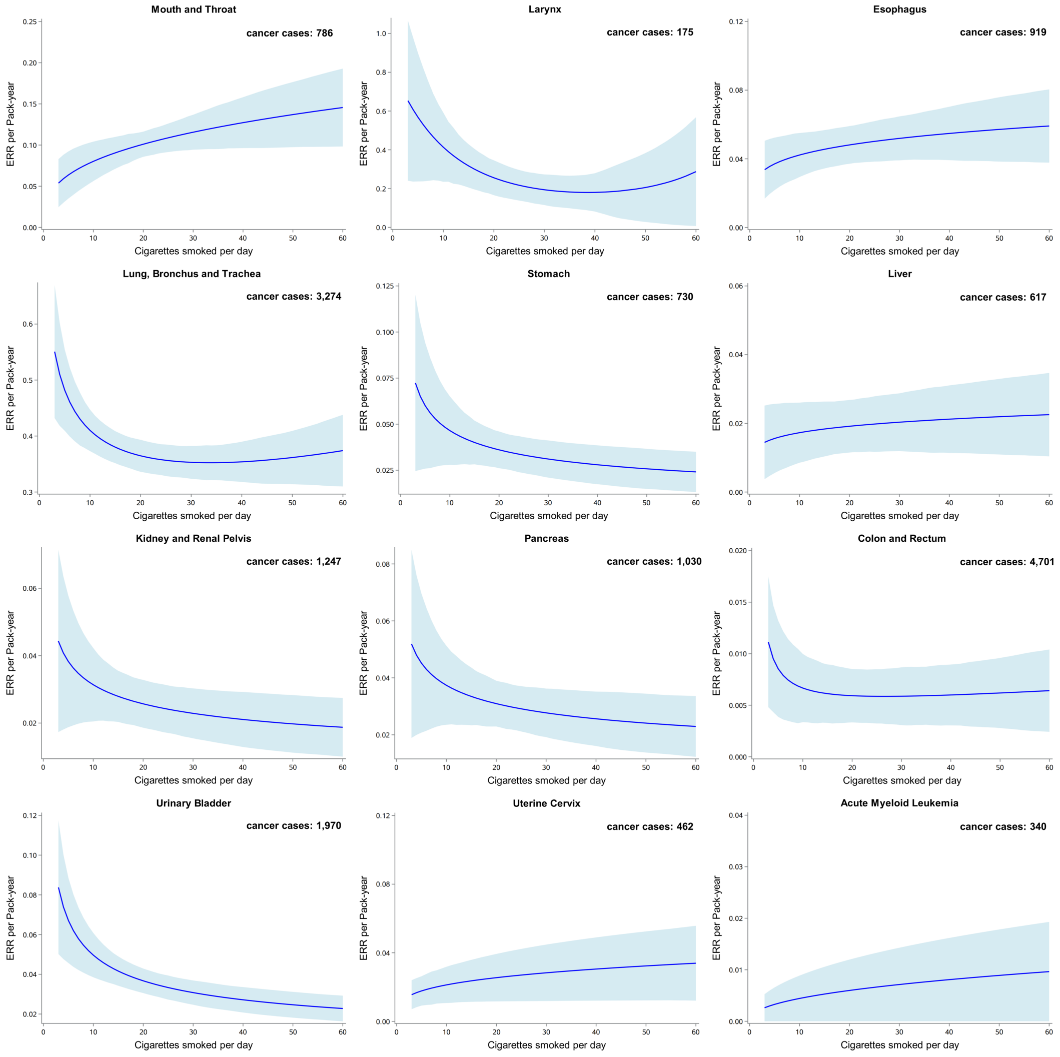

Bootstrapped 95% confidence intervals are based on 1,000 replications. The blue solid line represents the prediction from the function $g_1(n)$ in the model, with the area showing 95% confidence interval obtained from bootstrap analysis.

**Model adjustments:** age (<45, 45–50, 50–60, 60–65, ≥65, years), sex (men or women), ethnicity (White, Asian or Asian British, Black or Black British, Chinese, Mixed, other ethnic group, or unknown), BMI (<18.5, 18.5–25, 25–30, ≥30, or unknown, $kg/m^2$), socioeconomic status (low, medium, high, or unknown), alcohol consumption status (never, previous, current, or unknown), physical activity (regular physical activity, un-regular physical activity, or unknown).

**Abbreviations:** ERR, excess relative risk; BMI, body mass index.

**Figure S4.** The excess relative risk (ERR) for pan-cancers per pack-year of smoking by smoking intensity (adjusted for age, sex, BMI, ethnicity, SES, alcohol consumption status, healthy diet)

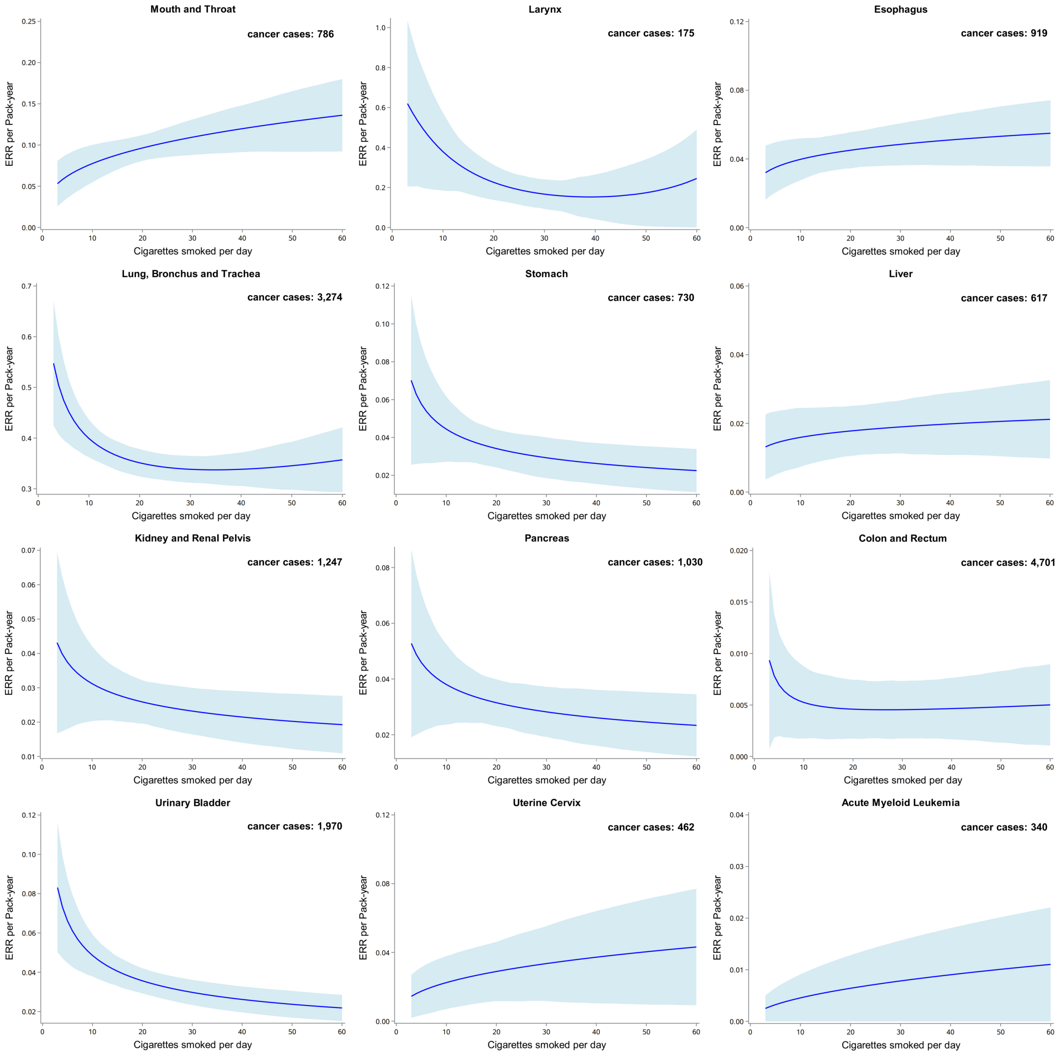

Bootstrapped 95% confidence intervals are based on 1,000 replications. The blue solid line represents the prediction from the function $g_1(n)$ in the model, with the area showing 95% confidence interval obtained from bootstrap analysis.
**Model adjustments:** age (<45, 45–50, 50–60, 60–65, ≥65, years), sex (men or women), ethnicity (White, Asian or Asian British, Black or Black British, Chinese, Mixed, other ethnic group, or unknown), BMI (<18.5, 18.5–25, 25–30, ≥30, or unknown, kg/m$^2$), socioeconomic status (low, medium, high, or unknown), and alcohol consumption status (never, previous, current, or unknown), healthy diet (healthy diet, unhealthy diet, unknown).
**Abbreviations:** ERR, excess relative risk; BMI, body mass index.

**Figure S5.** The excess relative risk (ERR) for pan-cancers per pack-year of smoking by smoking intensity (adjusted for age, sex, BMI, ethnicity, SES, alcohol consumption status, physical activity, healthy diet)

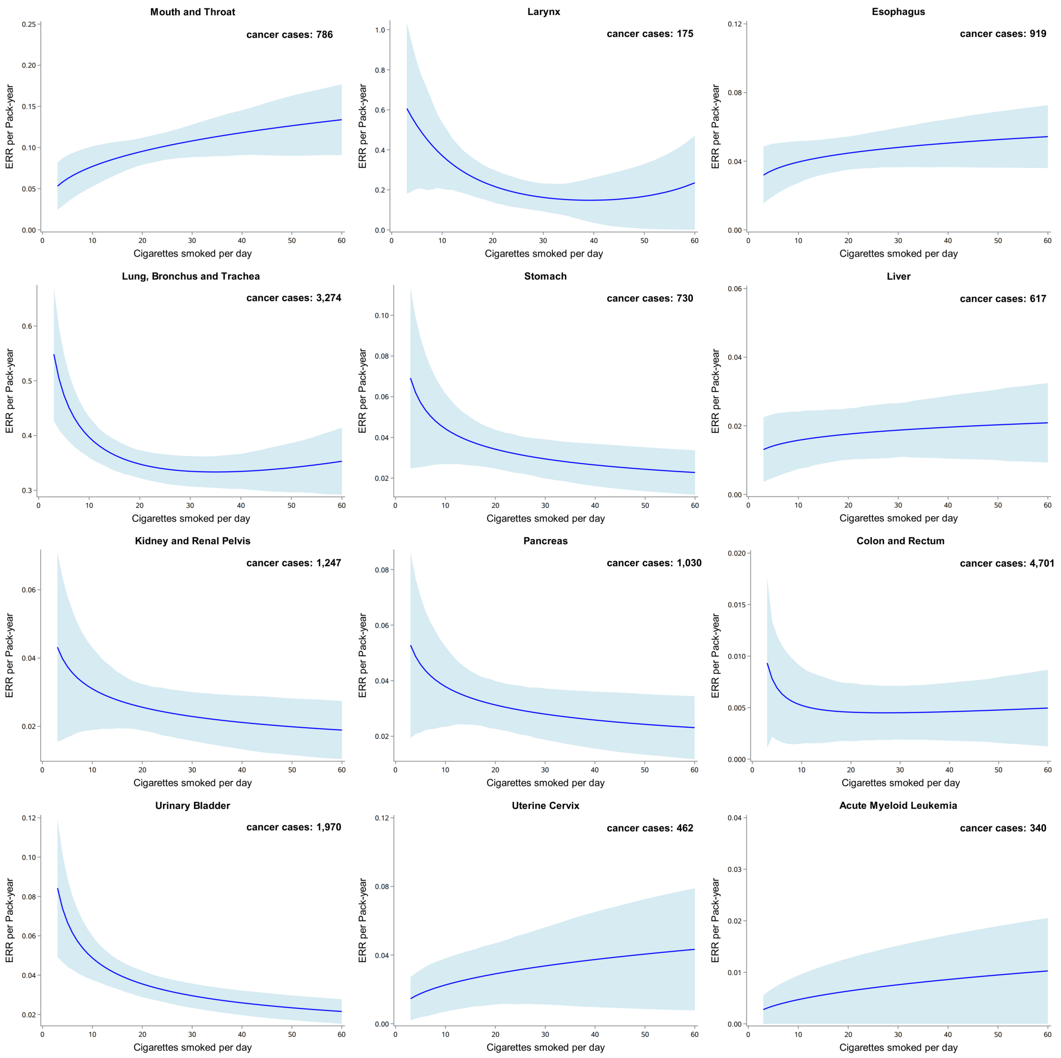

Bootstrapped 95% confidence intervals are based on 1,000 replications. The blue solid line represents the prediction from the function $g_1(n)$ in the model, with the area showing 95% confidence interval obtained from bootstrap analysis.

**Model adjustments:** age (<45, 45–50, 50–60, 60–65, ≥65, years), sex (men or women), ethnicity (White, Asian or Asian British, Black or Black British, Chinese, Mixed, other ethnic group, or unknown), BMI (<18.5, 18.5–25, 25–30, ≥30, or unknown, kg/m$^2$), socioeconomic status (low, medium, high, or unknown), and alcohol consumption status (never, previous, current, or unknown), physical activity (regular physical activity, un-regular physical activity, or unknown), and healthy diet (healthy diet, unhealthy diet, unknown).

**Abbreviations:** ERR, excess relative risk; BMI, body mass index.

**Figure S6.** Mean ERRs obtained based on main adjustment model across pan cancers

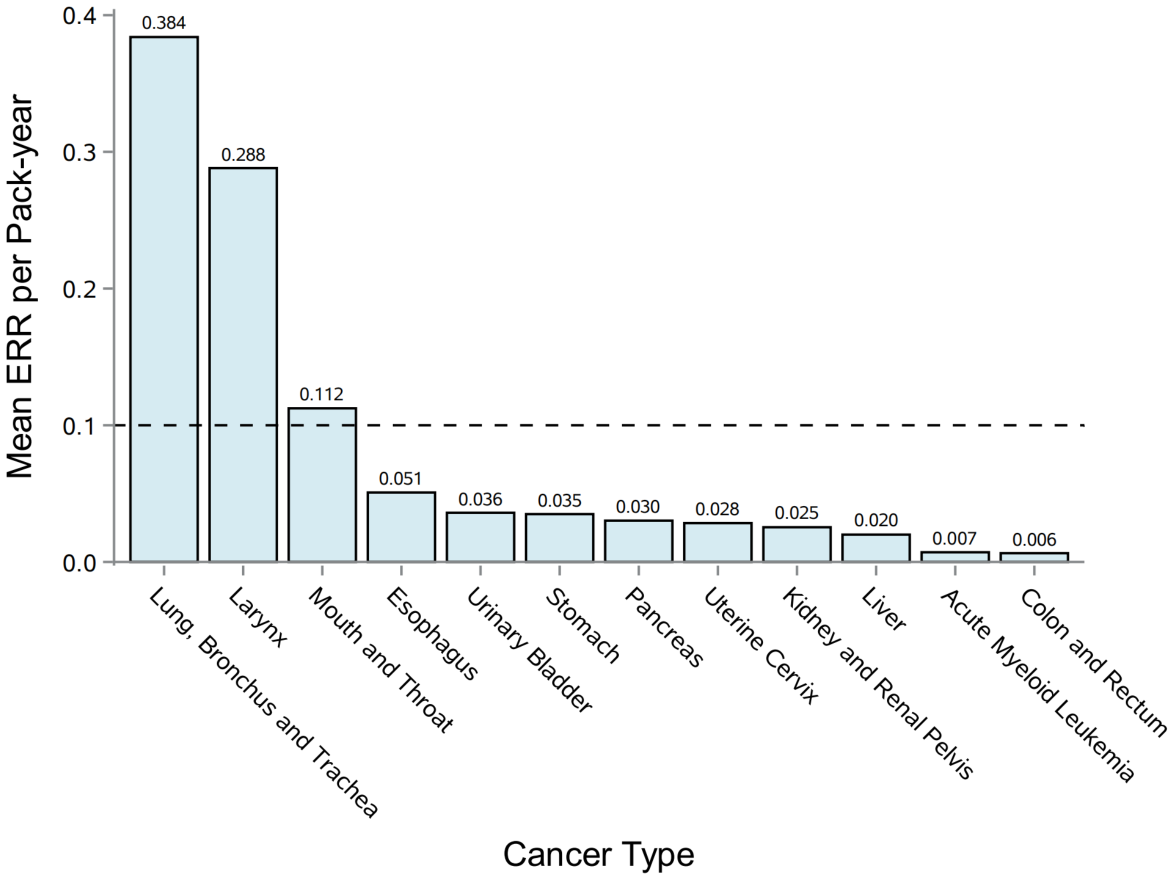

**Abbreviations:** ERR, excess relative risk.

**Figure S7.** The excess relative risk (ERR) for pan-cancers per pack-year of smoking by time since smoking cessation (without adjustments)

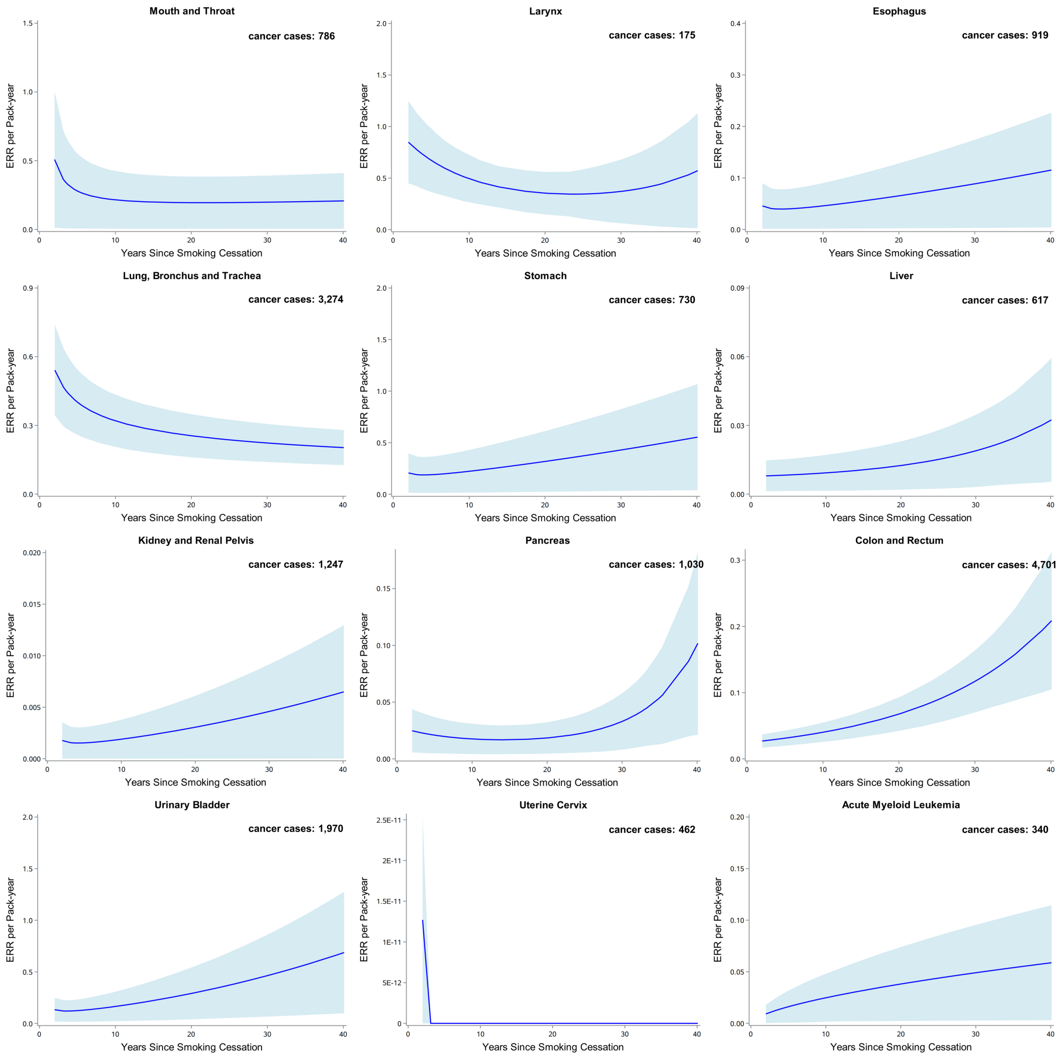

Bootstrapped 95% confidence intervals are based on 1,000 replications. The blue solid line represents the prediction from the function $g_2(t)$ in the model, with the area showing 95% confidence interval obtained from bootstrap analysis. The study subjects include never, current, and previous smokers. See text for details on models.
**Model adjustments:** without adjustments.
**Abbreviations:** ERR, excess relative risk; BMI, body mass index.

**Figure S8.** The excess relative risk (ERR) for pan-cancers per pack-year of smoking by time since smoking cessation (adjusted for age, sex, BMI, ethnicity)

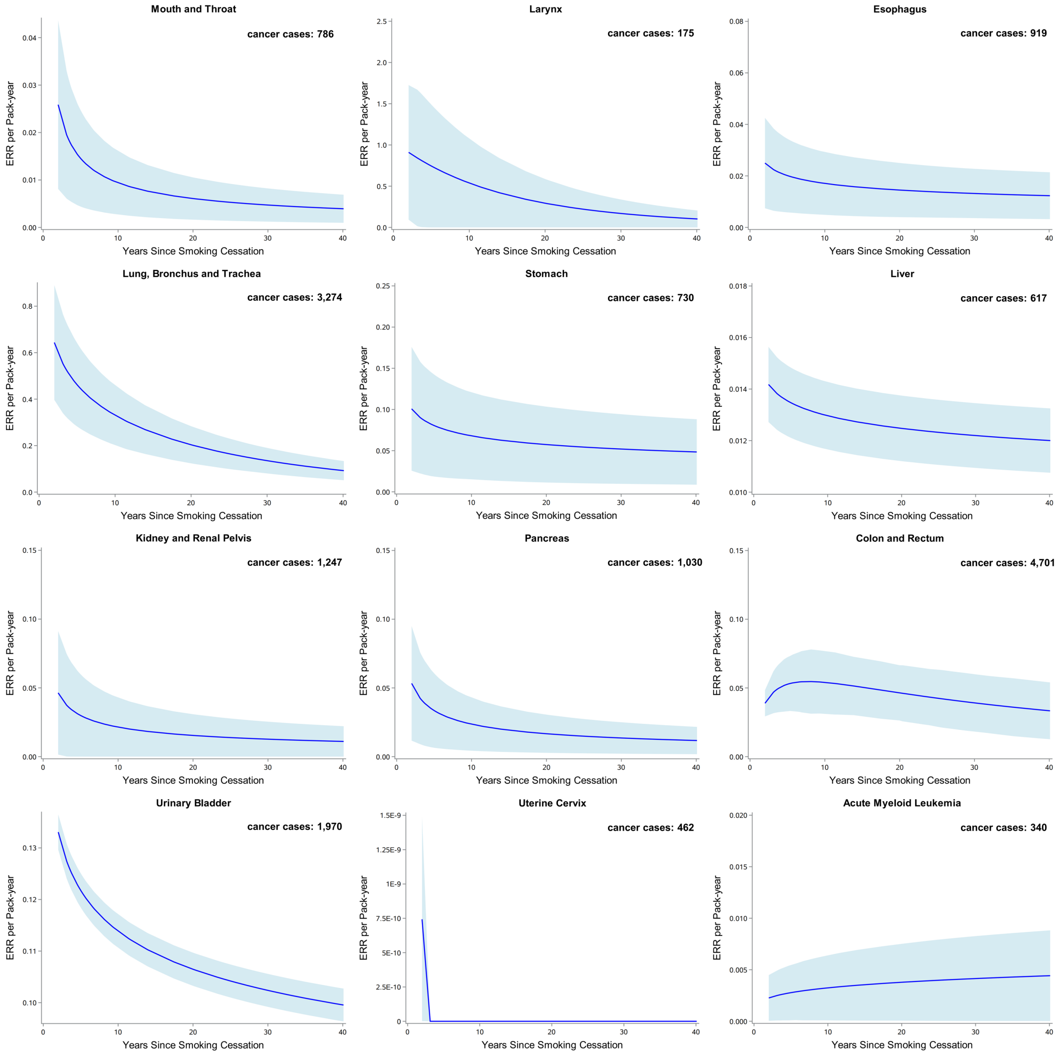

Bootstrapped 95% confidence intervals are based on 1,000 replications. The blue solid line represents the prediction from the function $g_2(t)$ in the model, with the area showing 95% confidence interval obtained from bootstrap analysis. The study subjects include never, current, and previous smokers. See text for details on models.
**Model adjustments:** age (<45, 45–50, 50–60, 60–65, ≥65, years), sex (men or women), ethnicity (White, Asian or Asian British, Black or Black British, Chinese, Mixed, other ethnic group, or unknown), BMI (<18.5, 18.5–25, 25–30, ≥30, or unknown, $kg/m^2$).
**Abbreviations:** ERR, excess relative risk; BMI, body mass index.

**Figure S9.** The excess relative risk (ERR) for pan-cancers per pack-year of smoking by time since smoking cessation (adjusted for age, sex, BMI, ethnicity, SES, alcohol consumption status, physical activity)

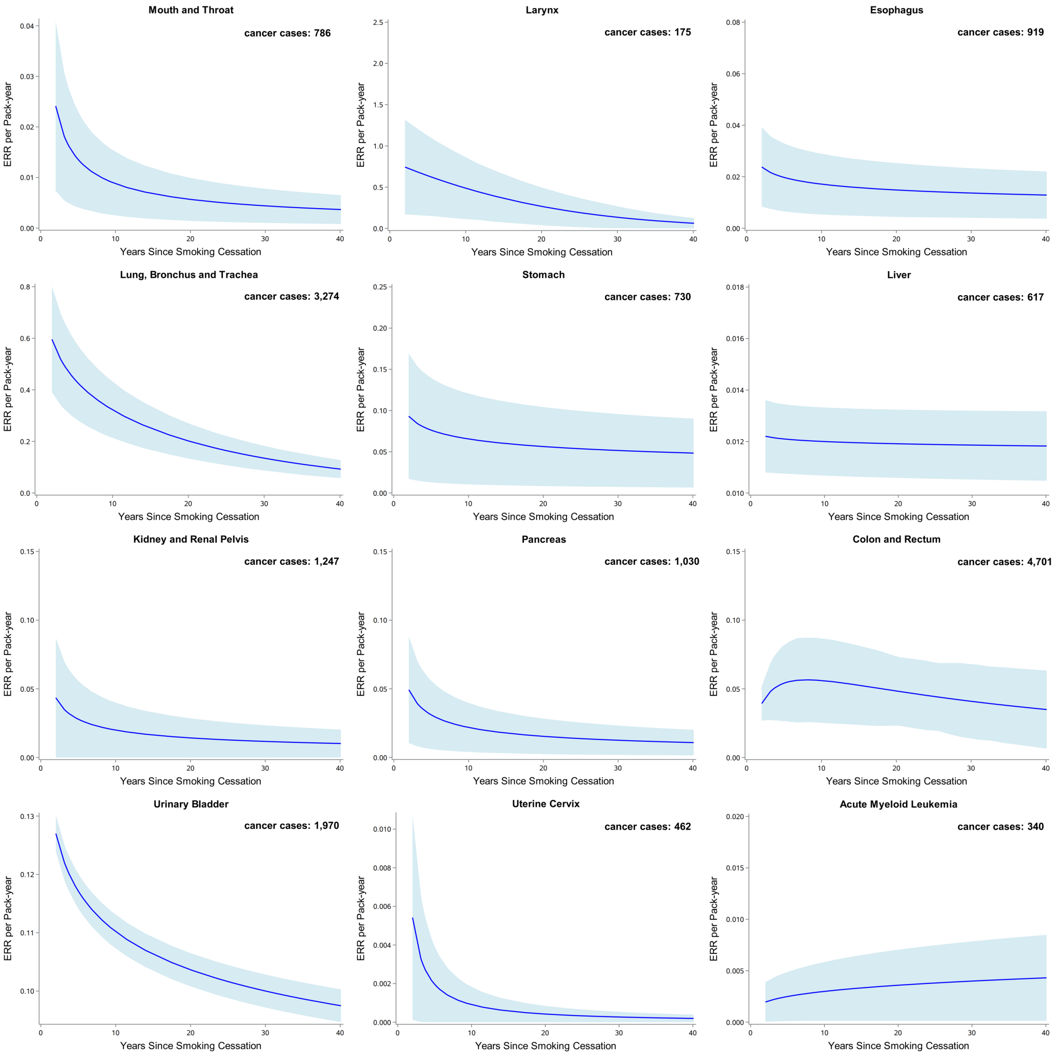

Bootstrapped 95% confidence intervals are based on 1,000 replications. The blue solid line represents the prediction from the function $g_2(t)$ in the model, with the area showing 95% confidence interval obtained from bootstrap analysis. The study subjects include never, current, and previous smokers. See text for details on models.
**Model adjustments:** age (<45, 45–50, 50–60, 60–65, ≥65, years), sex (men or women), ethnicity (White, Asian or Asian British, Black or Black British, Chinese, Mixed, other ethnic group, or unknown), BMI (<18.5, 18.5–25, 25–30, ≥30, or unknown, $kg/m^2$), socioeconomic status (low, medium, high, or unknown), alcohol consumption status (never, previous, current, or unknown), and physical activity (regular physical activity, un-regular physical activity, or unknown).
**Abbreviations:** ERR, excess relative risk; BMI, body mass index.

**Figure S10.** The excess relative risk (ERR) for pan-cancers per pack-year of smoking by time since smoking cessation (adjusted for age, sex, BMI, ethnicity, SES, alcohol consumption status, healthy diet)

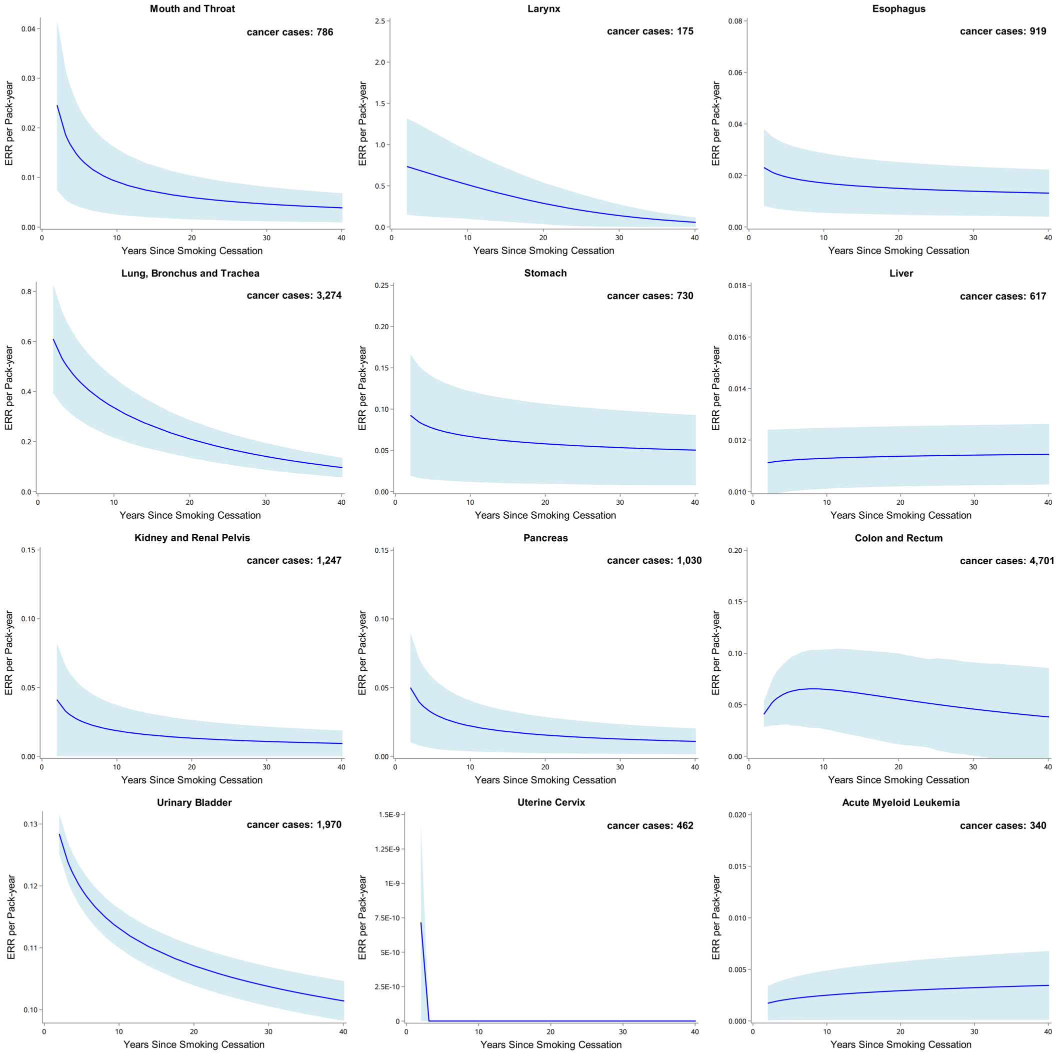

Bootstrapped 95% confidence intervals are based on 1,000 replications. The blue solid line represents the prediction from the function $g_2(t)$ in the model, with the area showing 95% confidence interval obtained from bootstrap analysis. The study subjects include never, current, and previous smokers. See text for details on models.
**Model adjustments:** age (<45, 45–50, 50–60, 60–65, ≥65, years), sex (men or women), ethnicity (White, Asian or Asian British, Black or Black British, Chinese, Mixed, other ethnic group, or unknown), BMI (<18.5, 18.5–25, 25–30, ≥30, or unknown, $kg/m^2$), socioeconomic status (low, medium, high, or unknown), alcohol consumption status (never, previous, current, or unknown), and healthy diet (healthy diet, unhealthy diet, unknown).
**Abbreviations:** ERR, excess relative risk; BMI, body mass index.

**Figure S11.** The excess relative risk (ERR) for pan-cancers per pack-year of smoking by time since smoking cessation (adjusted for age, sex, BMI, ethnicity, SES, alcohol consumption status, physical activity, healthy diet)

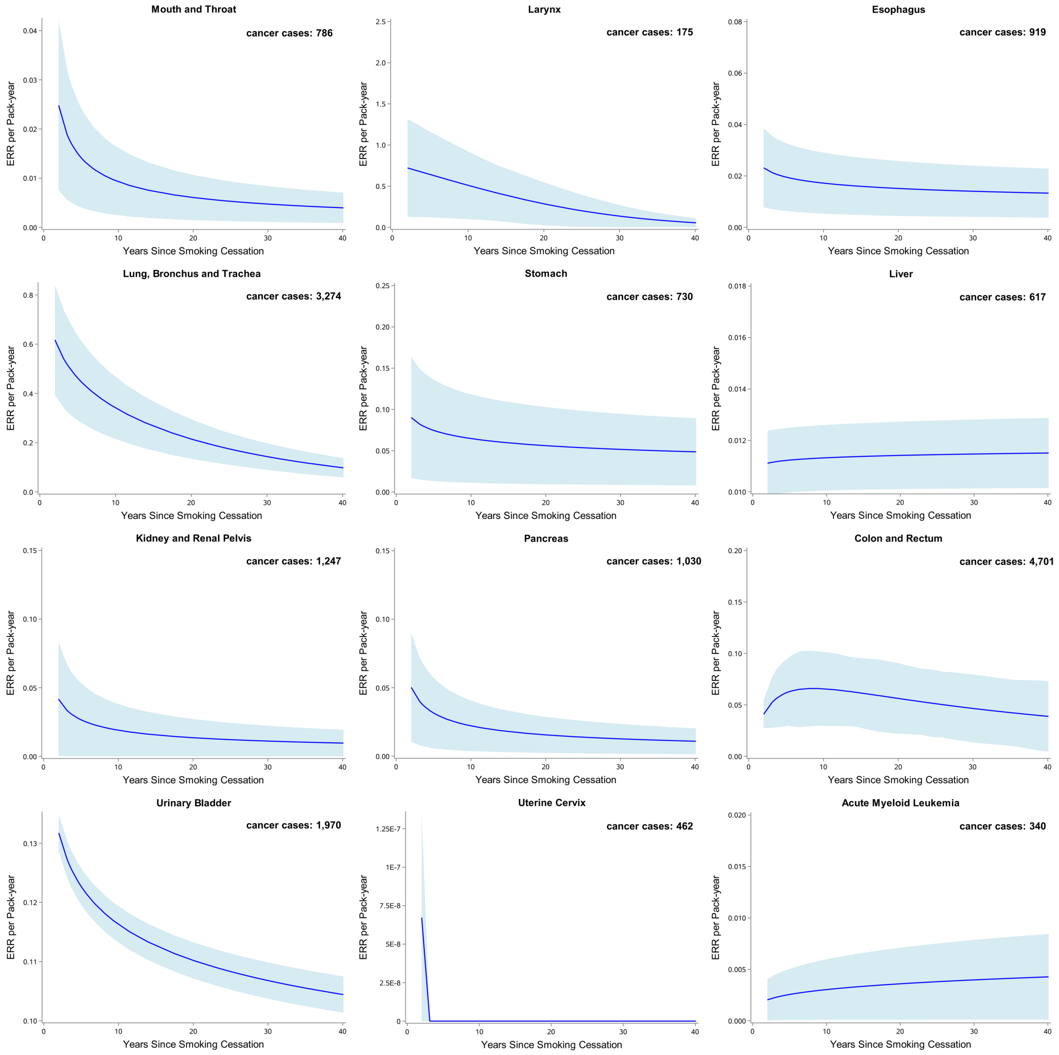

Bootstrapped 95% confidence intervals are based on 1,000 replications. The blue solid line represents the prediction from the function $g_2(t)$ in the model, with the area showing 95% confidence interval obtained from bootstrap analysis. The study subjects include never, current, and previous smokers. See text for details on models.
**Model adjustments:** age (<45, 45–50, 50–60, 60–65, ≥65, years), sex (men or women), ethnicity (White, Asian or Asian British, Black or Black British, Chinese, Mixed, other ethnic group, or unknown), BMI (<18.5, 18.5–25, 25–30, ≥30, or unknown, kg/m$^2$), socioeconomic status (low, medium, high, or unknown), alcohol consumption status (never, previous, current, or unknown), physical activity (regular physical activity, un-regular physical activity, or unknown), and healthy diet (healthy diet, unhealthy diet, unknown).
**Abbreviations:** ERR, excess relative risk; BMI, body mass index.

**Figure S12.** The excess relative risk (ERR) for pan-cancers per pack-year of smoking by smoking intensity (limiting subjects to ages 50–74 years at enrollment and regrouping the former smokers who quitted smoking within 5 years at baseline as current smokers)

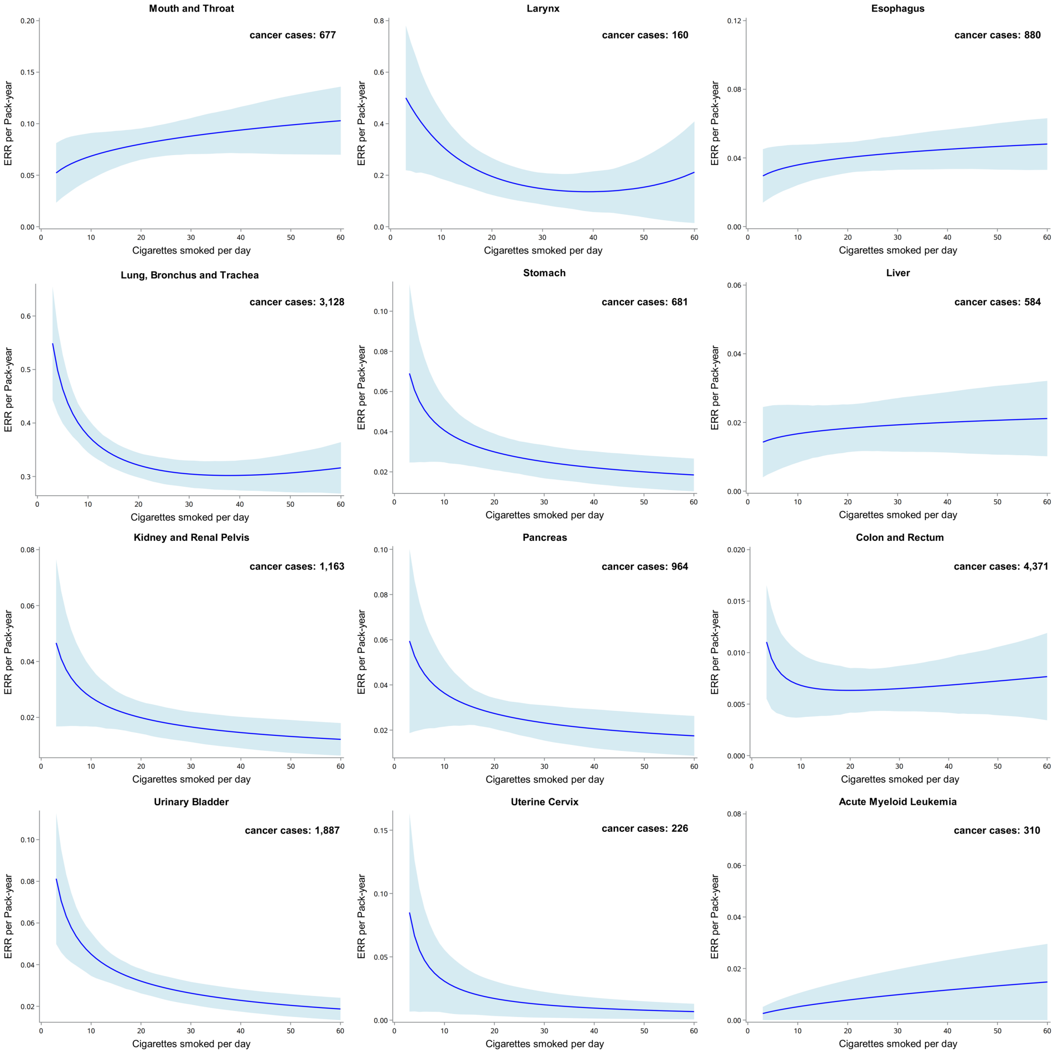

Bootstrapped 95% confidence intervals are based on 1,000 replications. The blue solid line represents the prediction from the function $g_1(n)$ in the model, with the area showing 95% confidence interval obtained from bootstrap analysis.

**Model adjustments:** age (50–60, 60–65, 65–74, years), sex (men or women), ethnicity (White, Asian or Asian British, Black or Black British, Chinese, Mixed, other ethnic group, or unknown), BMI (<18.5, 18.5–25, 25–30, ≥30, or unknown, kg/m$^2$), socioeconomic status (low, medium, high, or unknown), and alcohol consumption status (never, previous, current, or unknown).

**Abbreviations:** ERR, excess relative risk; BMI, body mass index.

**Figure S13.** The excess relative risk (ERR) for pan-cancers per pack-year of smoking by smoking intensity (excluding non-White individuals)

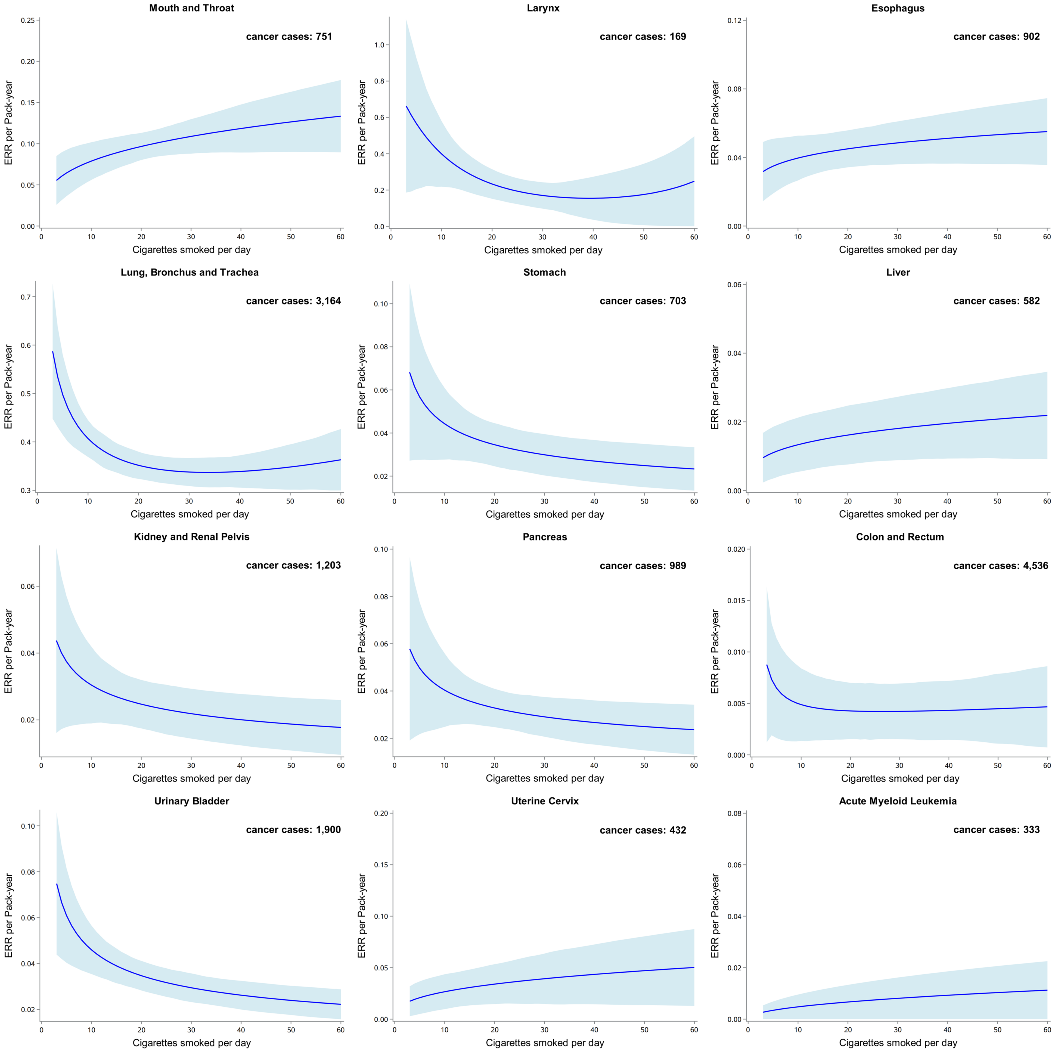

Bootstrapped 95% confidence intervals are based on 1,000 replications. The blue solid line represents the prediction from the function $g_1(n)$ in the model, with the area showing 95% confidence interval obtained from bootstrap analysis.

**Model adjustments:** age (<45, 45–50, 50–60, 60–65, ≥65, years), sex (men or women), BMI (<18.5, 18.5–25, 25–30, ≥30, or unknown, kg/m$^2$), socioeconomic status (low, medium, high, or unknown), and alcohol consumption status (never, previous, current, or unknown).

**Abbreviations:** ERR, excess relative risk; BMI, body mass index.

**Figure S14.** The excess relative risk (ERR) for pan-cancers per pack-year of smoking by smoking intensity (excluding incident cases of cancer occurring during the first year of follow-up)

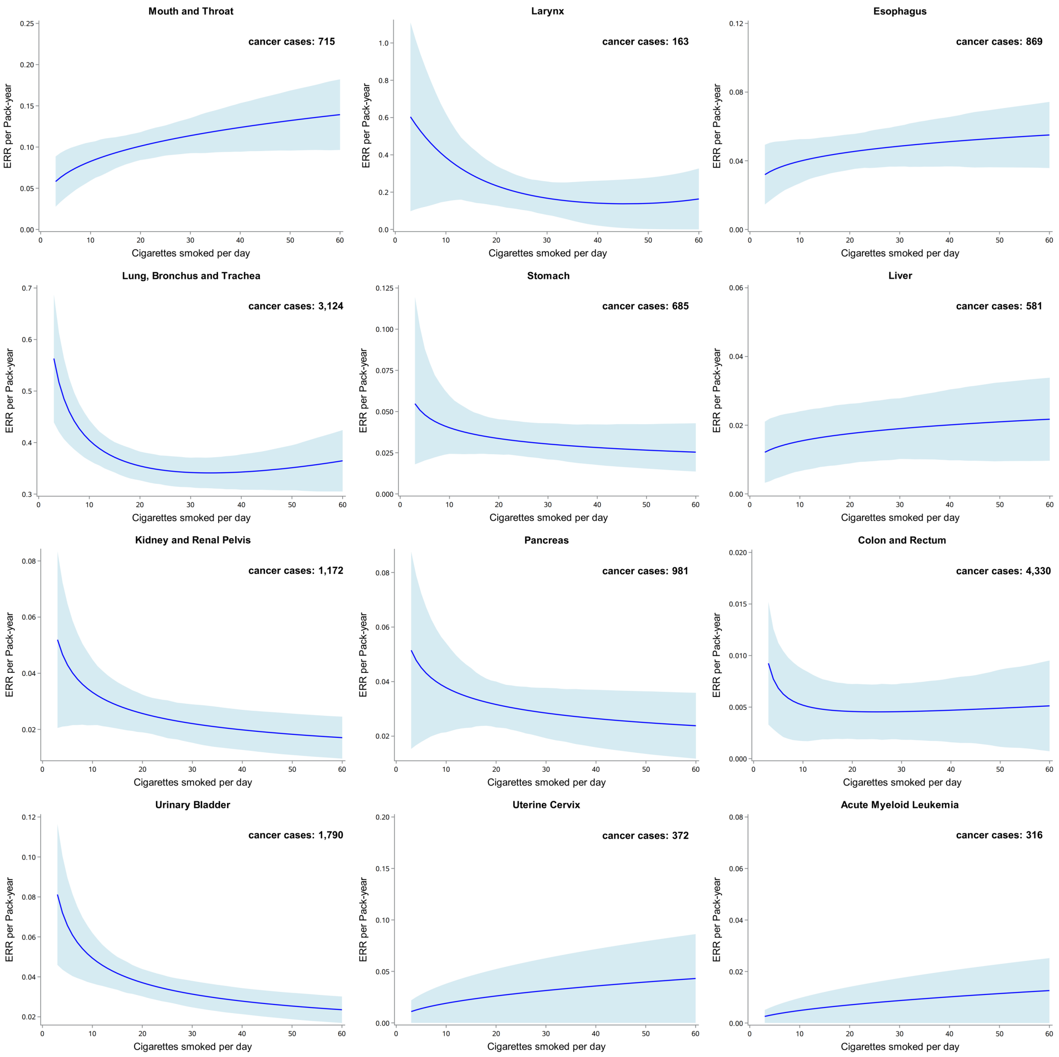

Bootstrapped 95% confidence intervals are based on 1,000 replications. The blue solid line represents the prediction from the function $g_1(n)$ in the model, with the area showing 95% confidence interval obtained from bootstrap analysis.

**Model adjustments:** age (<45, 45–50, 50–60, 60–65, ≥65, years), sex (men or women), ethnicity (White, Asian or Asian British, Black or Black British, Chinese, Mixed, other ethnic group, or unknown), BMI (<18.5, 18.5–25, 25–30, ≥30, or unknown, kg/m$^2$), socioeconomic status (low, medium, high, or unknown), and alcohol consumption status (never, previous, current, or unknown).

**Abbreviations:** ERR, excess relative risk; BMI, body mass index.

**Figure S15.** The excess relative risk (ERR) for pan-cancers per pack-year of smoking by smoking intensity (excluding the never smokers who reported with passive smoking)

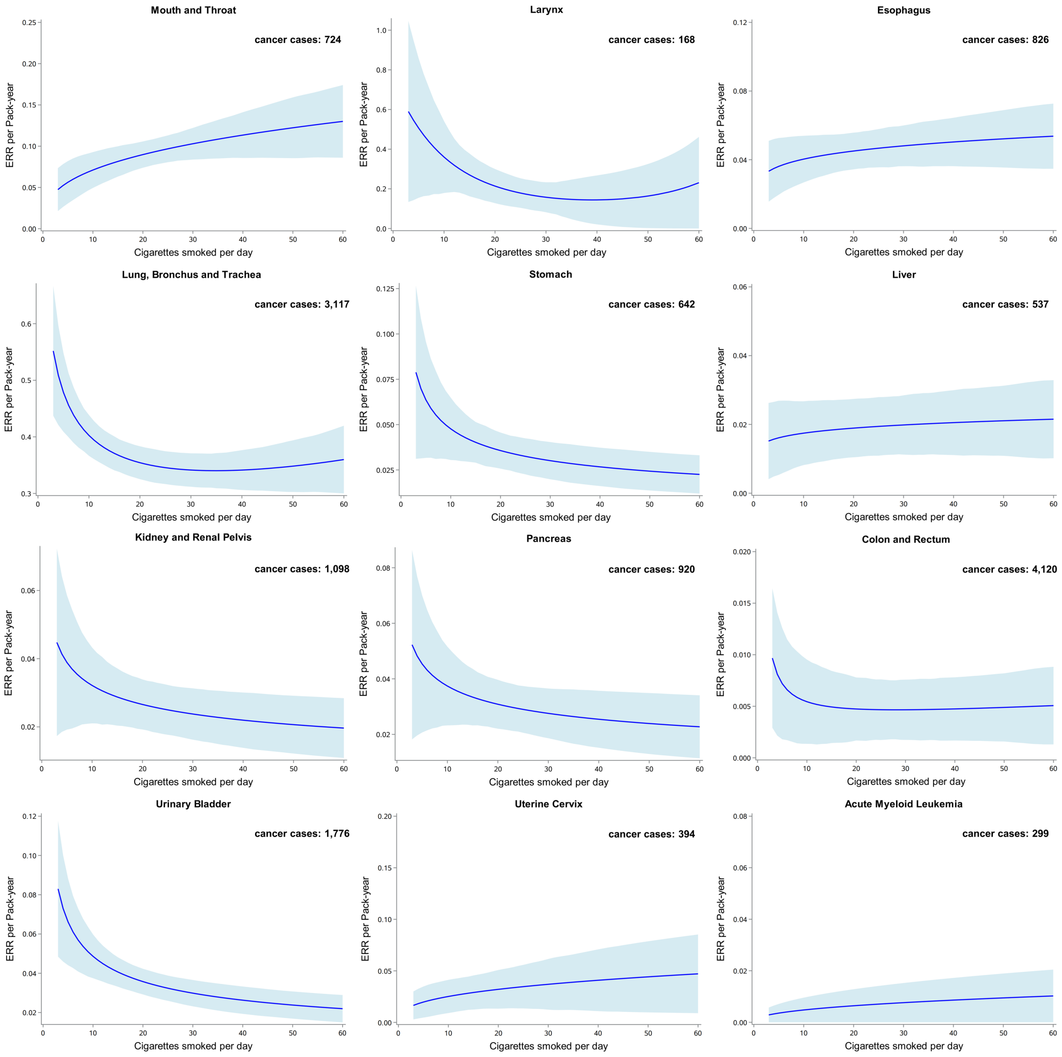

Bootstrapped 95% confidence intervals are based on 1,000 replications. The blue solid line represents the prediction from the function $g_1(n)$ in the model, with the area showing 95% confidence interval obtained from bootstrap analysis.

**Model adjustments:** age (<45, 45–50, 50–60, 60–65, ≥65, years), sex (men or women), ethnicity (White, Asian or Asian British, Black or Black British, Chinese, Mixed, other ethnic group, or unknown), BMI (<18.5, 18.5–25, 25–30, ≥30, or unknown, $kg/m^2$), socioeconomic status (low, medium, high, or unknown), and alcohol consumption status (never, previous, current, or unknown).

**Abbreviations:** ERR, excess relative risk; BMI, body mass index.

**Figure S16.** The excess relative risk (ERR) for pan-cancers per pack-year of smoking by smoking intensity (considering the influence of frequency and amount of current alcohol consumption)

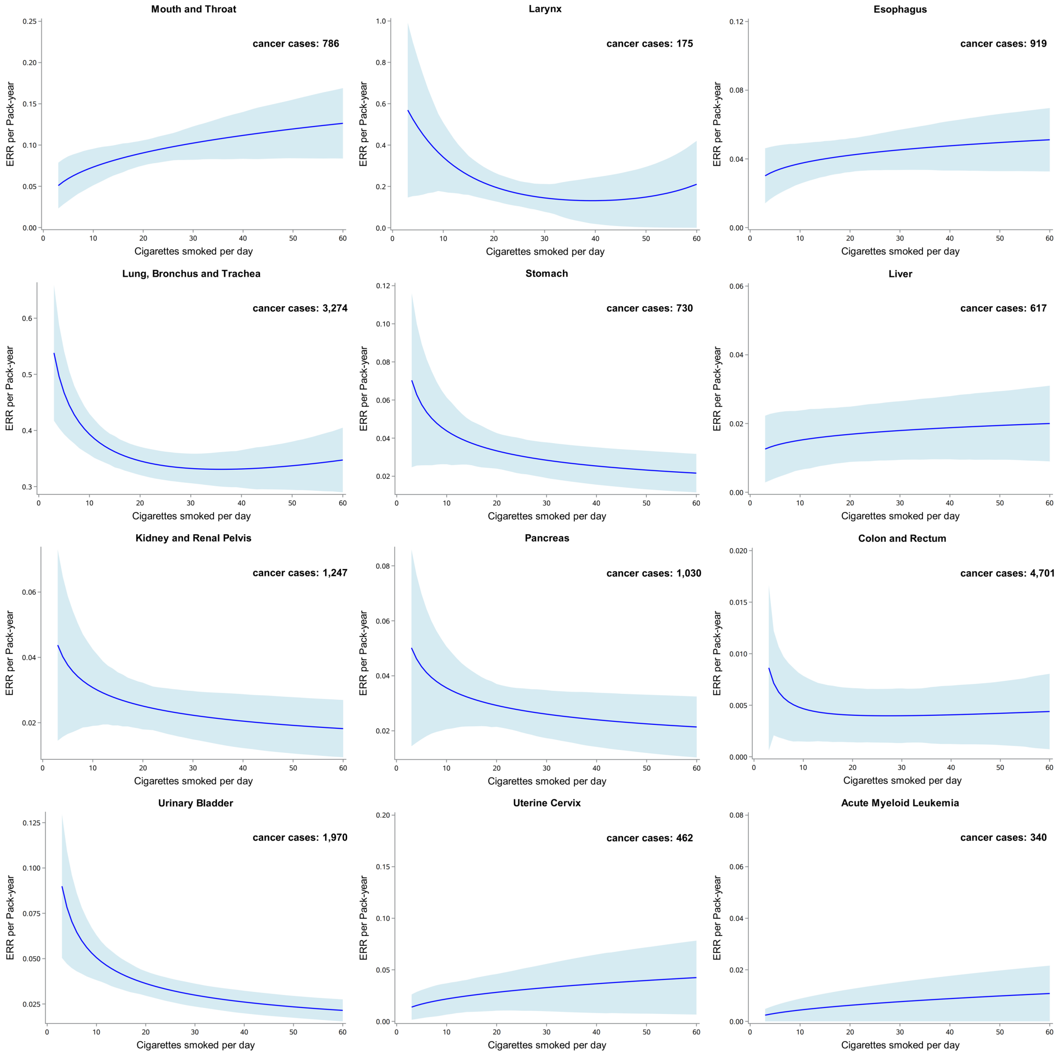

Bootstrapped 95% confidence intervals are based on 1,000 replications. The blue solid line represents the prediction from the function $g_1(n)$ in the model, with the area showing 95% confidence interval obtained from bootstrap analysis.

**Model adjustments:** age (<45, 45–50, 50–60, 60–65, ≥65, years), sex (men or women), ethnicity (White, Asian or Asian British, Black or Black British, Chinese, Mixed, other ethnic group, or unknown), BMI (<18.5, 18.5–25, 25–30, ≥30, or unknown, kg/m$^2$), socioeconomic status (low, medium, high, or unknown), alcohol consumption status (never, previous, current, or unknown), and alcohol frequency and amount.

**Abbreviations:** ERR, excess relative risk; BMI, body mass index.

**Figure S17.** The excess relative risk (ERR) for pan-cancers per pack-year of smoking by time since smoking cessation. (limiting subjects to ages 50–74 years at enrollment and regrouping the former smokers who quitted smoking within 5 years at baseline as current smokers)

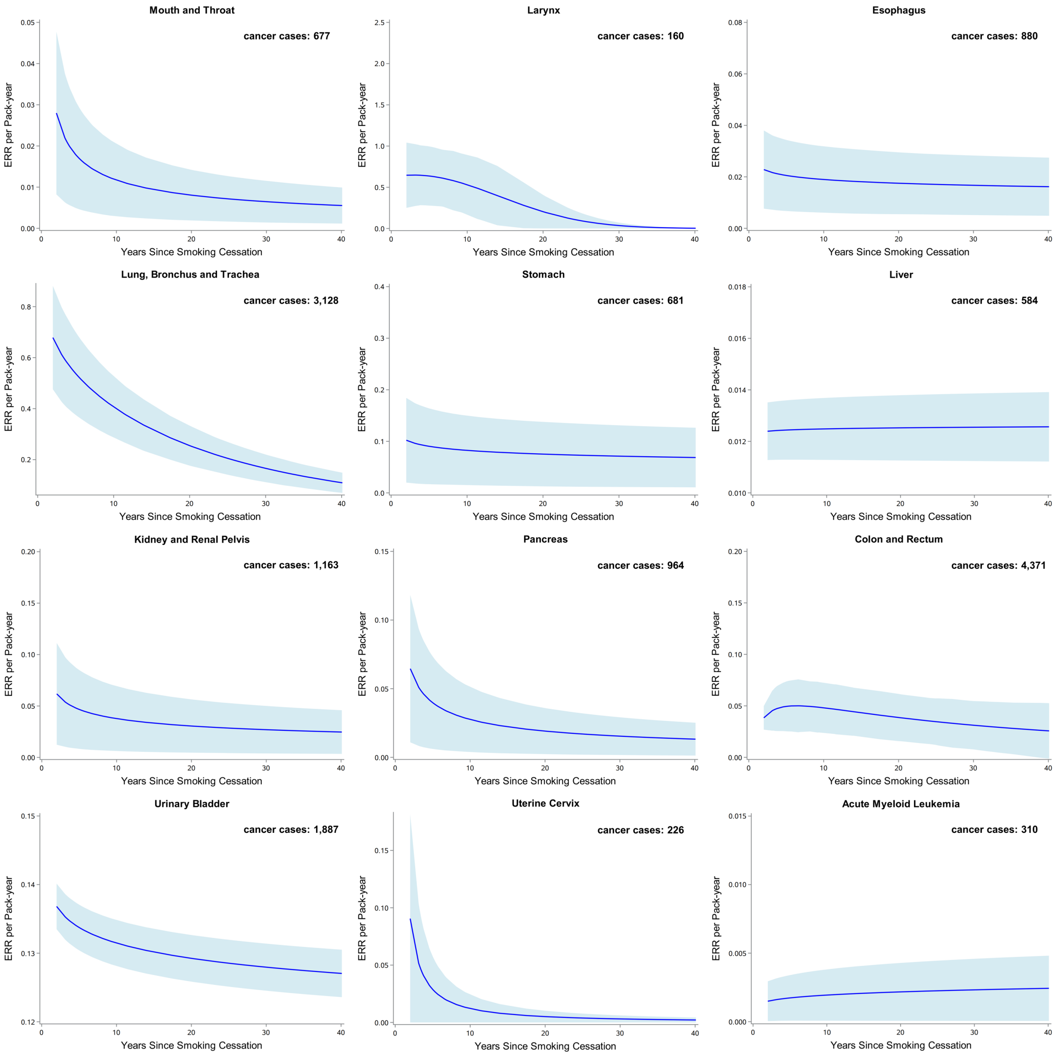

Bootstrapped 95% confidence intervals are based on 1,000 replications. The blue solid line represents the prediction from the function $g_2(t)$ in the model, with the area showing 95% confidence interval obtained from bootstrap analysis. The study subjects include never, current, and previous smokers. See text for details on models.
**Model adjustments:** age (50–60, 60–65, 65–74, years), sex (men or women), ethnicity (White, Asian or Asian British, Black or Black British, Chinese, Mixed, other ethnic group, or unknown), BMI (<18.5, 18.5–25, 25–30, ≥30, or unknown, kg/m$^2$), socioeconomic status (low, medium, high, or unknown), and alcohol consumption status (never, previous, current, or unknown).
**Abbreviations:** ERR, excess relative risk; BMI, body mass index.

**Figure S18.** The excess relative risk (ERR) for pan-cancers per pack-year of smoking by time since smoking cessation (excluding non-White individuals)

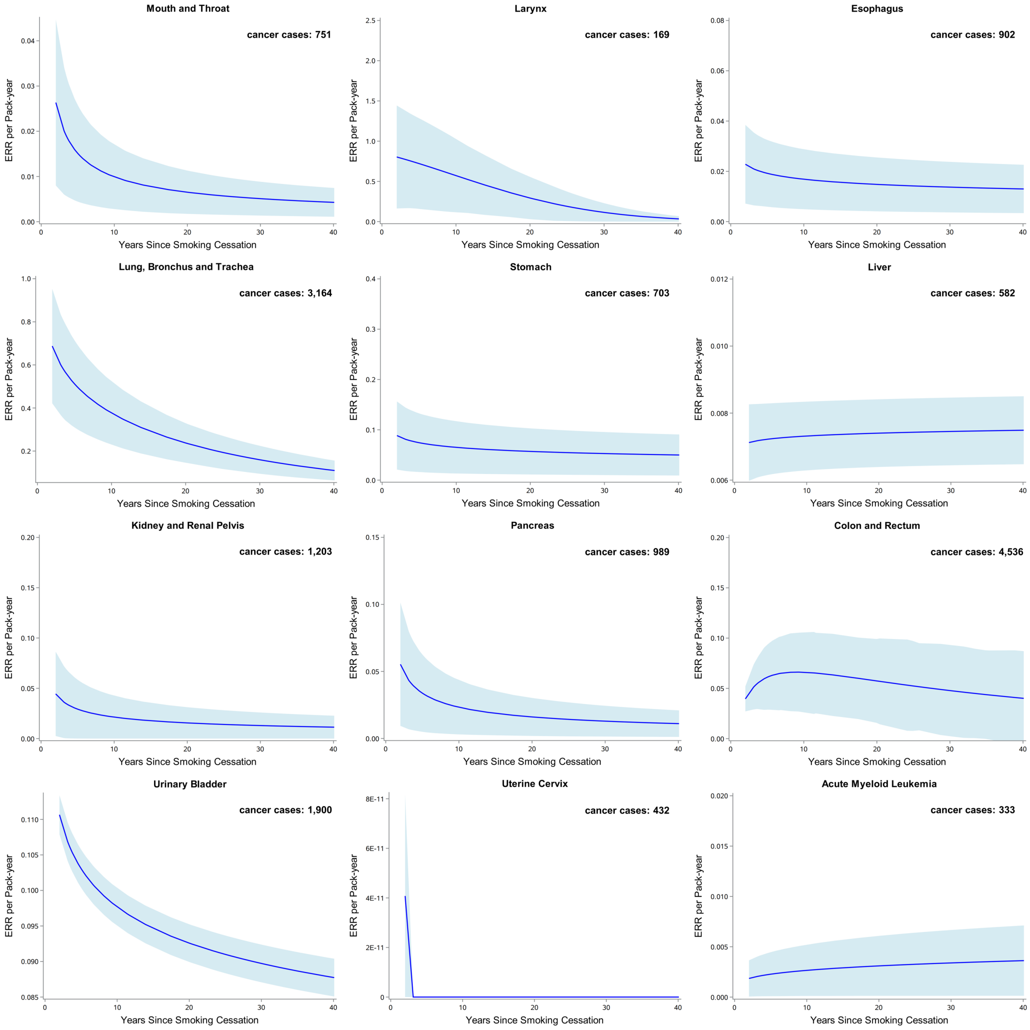

Bootstrapped 95% confidence intervals are based on 1,000 replications. The blue solid line represents the prediction from the function $g_2(t)$ in the model, with the area showing 95% confidence interval obtained from bootstrap analysis. The study subjects include never, current, and previous smokers. See text for details on models.
**Model adjustments:** age (<45, 45–50, 50–60, 60–65, ≥65, years), sex (men or women), BMI (<18.5, 18.5–25, 25–30, ≥30, or unknown, kg/m$^2$), socioeconomic status (low, medium, high, or unknown), and alcohol consumption status (never, previous, current, or unknown).
**Abbreviations:** ERR, excess relative risk; BMI, body mass index.

**Figure S19.** The excess relative risk (ERR) for pan-cancers per pack-year of smoking by time since smoking cessation (excluding incident cases of cancer occurring during the first year of follow-up)

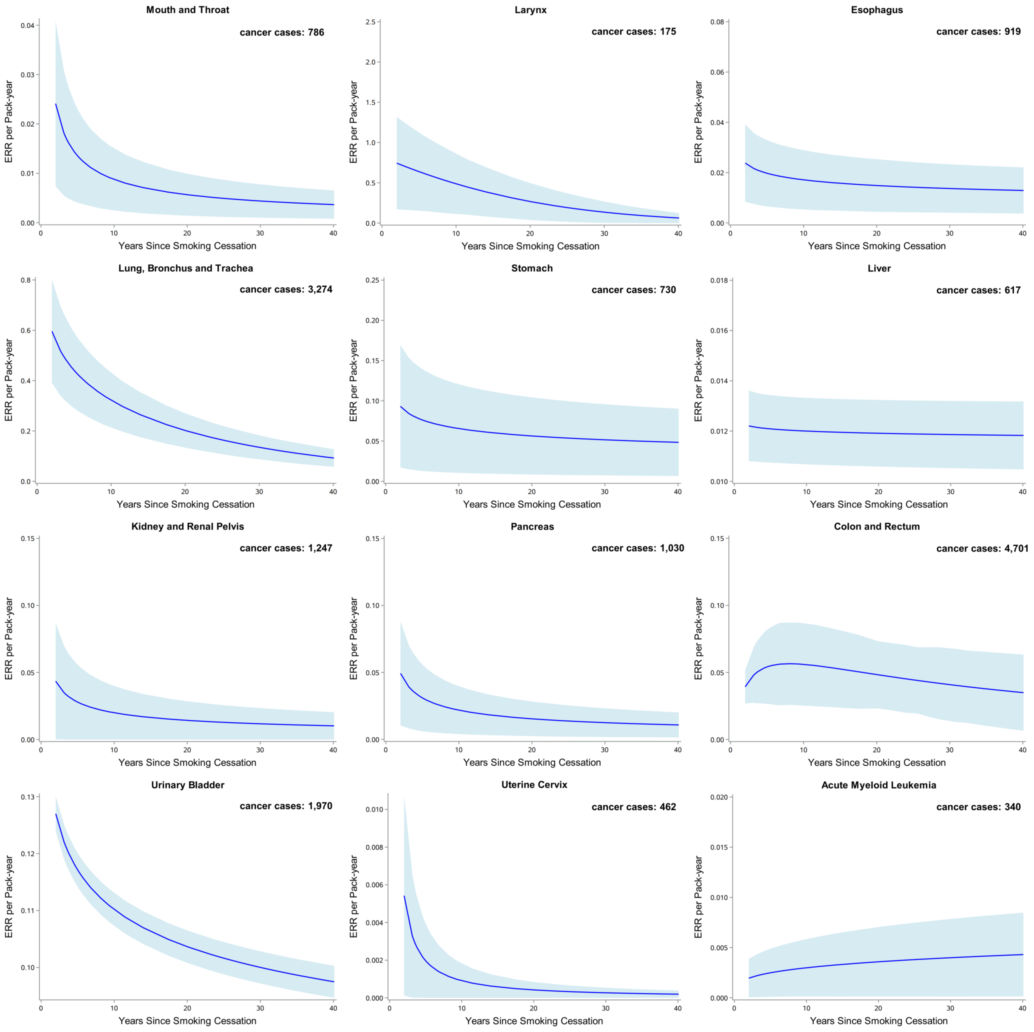

Bootstrapped 95% confidence intervals are based on 1,000 replications. The blue solid line represents the prediction from the function $g_2(t)$ in the model, with the area showing 95% confidence interval obtained from bootstrap analysis. The study subjects include never, current, and previous smokers. See text for details on models.
**Model adjustments:** age (<45, 45–50, 50–60, 60–65, ≥65, years), sex (men or women), ethnicity (White, Asian or Asian British, Black or Black British, Chinese, Mixed, other ethnic group, or unknown), BMI (<18.5, 18.5–25, 25–30, ≥30, or unknown, kg/m$^2$), socioeconomic status (low, medium, high, or unknown), and alcohol consumption status (never, previous, current, or unknown).
**Abbreviations:** ERR, excess relative risk; BMI, body mass index.

**Figure S20.** The excess relative risk (ERR) for pan-cancers per pack-year of smoking by time since smoking cessation (excluding the never smokers who reported with passive smoking)

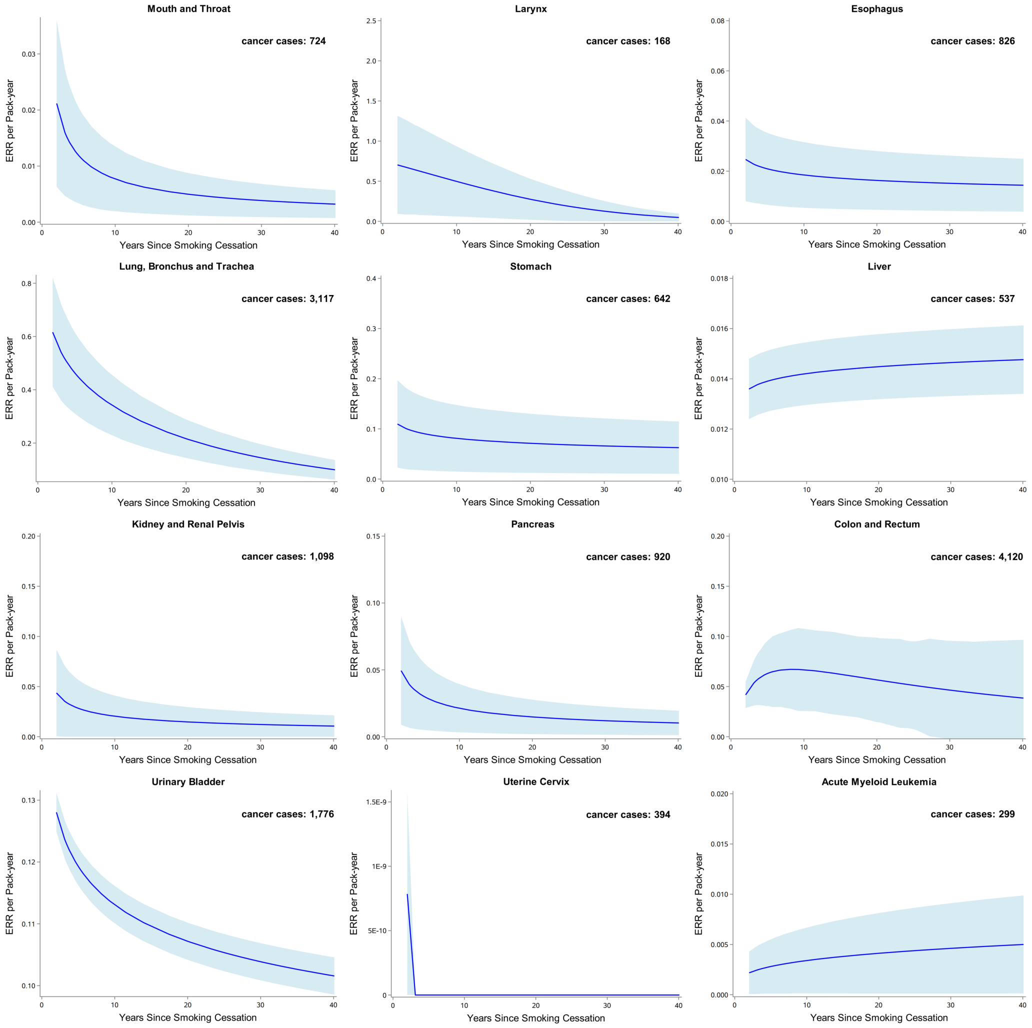

Bootstrapped 95% confidence intervals are based on 1,000 replications. The blue solid line represents the prediction from the function $g_2(t)$ in the model, with the area showing 95% confidence interval obtained from bootstrap analysis. The study subjects include never, current, and previous smokers. See text for details on models.
**Model adjustments:** age (<45, 45–50, 50–60, 60–65, ≥65, years), sex (men or women), ethnicity (White, Asian or Asian British, Black or Black British, Chinese, Mixed, other ethnic group, or unknown), BMI (<18.5, 18.5–25, 25–30, ≥30, or unknown, $kg/m^2$), socioeconomic status (low, medium, high, or unknown), and alcohol consumption status (never, previous, current, or unknown).
**Abbreviations:** ERR, excess relative risk; BMI, body mass index.

**Figure S21.** The excess relative risk (ERR) for pan-cancers per pack-year of smoking by time since smoking cessation (considering the influence of frequency and amount of current alcohol consumption)

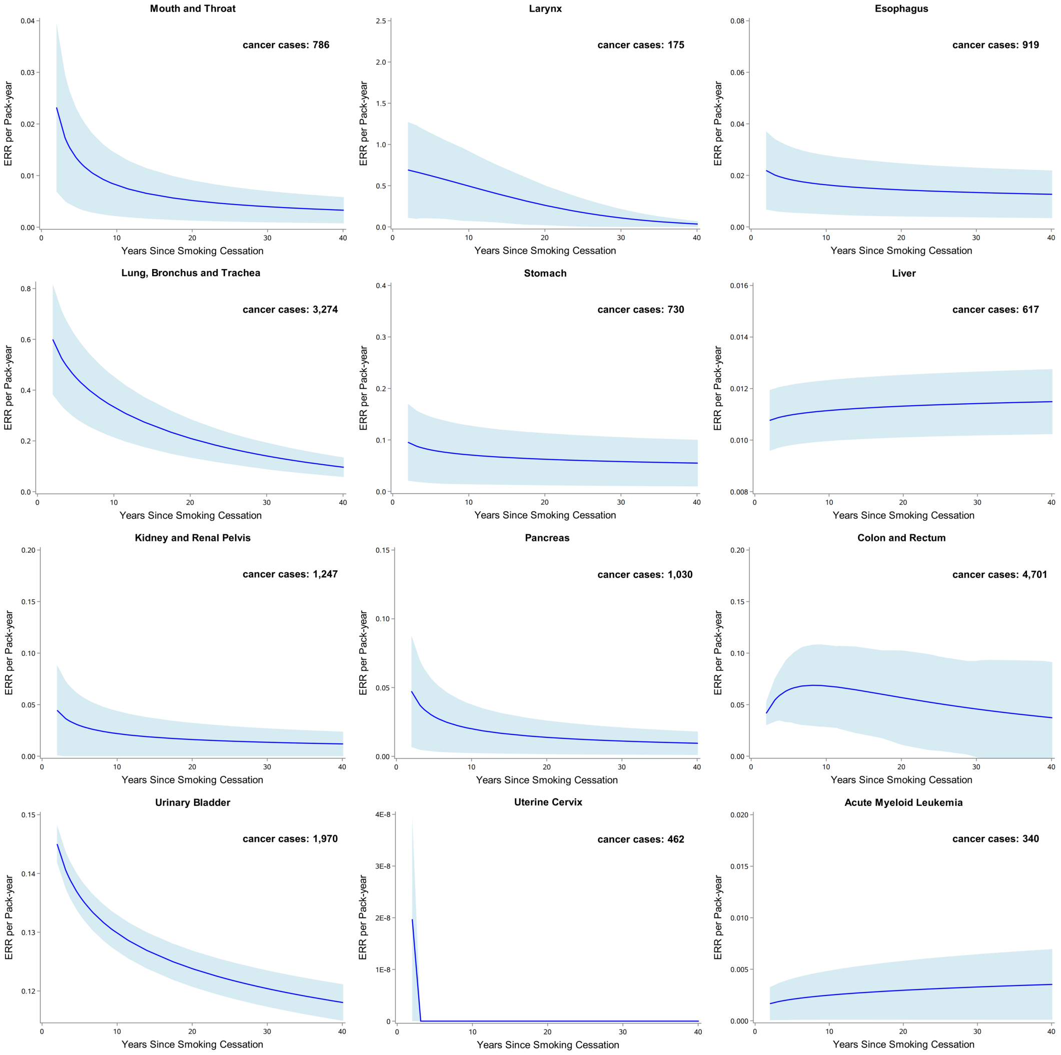

Bootstrapped 95% confidence intervals are based on 1,000 replications. The blue solid line represents the prediction from the function $g_2(t)$ in the model, with the area showing 95% confidence interval obtained from bootstrap analysis. The study subjects include never, current, and previous smokers. See text for details on models.
**Model adjustments:** age (<45, 45–50, 50–60, 60–65, ≥65, years), sex (men or women), ethnicity (White, Asian or Asian British, Black or Black British, Chinese, Mixed, other ethnic group, or unknown), BMI (<18.5, 18.5–25, 25–30, ≥30, or unknown, kg/m$^2$), socioeconomic status (low, medium, high, or unknown), alcohol consumption status (never, previous, current, or unknown), and alcohol frequency and amount.
**Abbreviations:** ERR, excess relative risk; BMI, body mass index.
